# Integrating Gut Microbiota and Host Immune Markers for Highly Accurate Diagnosis of *Clostridioides difficile* Infection

**DOI:** 10.1101/2020.04.27.20081653

**Authors:** Shanlin Ke, Nira R. Pollock, Xu-Wen Wang, Xinhua Chen, Kaitlyn Daugherty, Qianyun Lin, Hua Xu, Kevin W. Garey, Anne J. Gonzales-Luna, Ciarán P. Kelly, Yang-Yu Liu

**Author notes:** To whom correspondence should be addressed: Y.-Y.L. or C.P.K.

## Abstract

Exposure to *Clostridioides difficile* can result in asymptomatic carriage or infection with symptoms ranging from mild diarrhea to fulminant pseudomembranous colitis. A reliable diagnostic approach for *C. difficile* infection (CDI) remains controversial. Accurate diagnosis is paramount not only for patient management but also for epidemiology and disease research. Here, we characterized gut microbial compositions and a broad panel of innate and adaptive immunological markers in 243 well-characterized human subjects, who were divided into four phenotype groups: CDI, Asymptomatic Carriage, Non-CDI Diarrhea, and Control. We found that CDI is associated with alteration of many different aspects of the gut microbiota, including overall microbial diversity and microbial association networks. We demonstrated that incorporating both gut microbiome and host immune marker data into classification models can better distinguish CDI from other groups than can either type of data alone. Our classification models display robust diagnostic performance to differentiate CDI from Asymptomatic carriage (AUC~0.916), Non-CDI Diarrhea (AUC~0.917), or Non-CDI that combines all other three groups (AUC~0.929). Finally, we performed symbolic classification using selected features to derive simple mathematic formulas for highly accurate CDI diagnosis. Overall, this study provides evidence supporting important roles of gut microbiota and host immune markers in CDI diagnosis, which may also inform the design of future therapeutic strategies.

**One Sentence Summary:** Incorporating both gut microbiome and host immune marker data into classification models can better distinguish CDI from other groups than can either type of data alone.

## INTRODUCTION

*Clostridioides difficile* infection (CDI) is the most common cause of healthcare–associated infection and an important cause of morbidity and mortality among hospitalized patients^1–3^. Current treatment strategies for CDI, including vancomycin, metronidazole and fidaxomicin, have inconsistent cure rates and treatment failure or CDI recurrence may occur in approximately one third of cases^4,5^. Antibiotic exposure is considered the most important factor predisposing patients to CDI^6,7^. In fact, treatments with antibiotics have a tremendous impact on the composition and functionality of the gut microbiota, and accordingly are associated with reduced colonization resistance against pathogens such as *C. difficile*^8–10^. A distinct microbial community structure has been reported to be associated with CDI in human cohorts and animal models^11,12^. Characterization of the microbial features in individuals with different *C. difficile* infection/colonization status is an essential step in understanding the role of the gut microbiome in the development of CDI.

The pathophysiology of CDI is mainly associated with the production of two exotoxins, toxin A (TcdA) and toxin B (TcdB)^13^. TcdA and TcdB act on intestinal epithelial cells, inducing pro-inflammatory cytokines, loss of tight junctions, cell detachment and an impaired mucosal barrier^14–16^. The innate and adaptive immune responses to CDI play crucial roles in disease onset, expression, severity, progression, and overall prognosis^17,18^. The innate immune defense mechanisms against *C. difficile* and its toxins include the commensal intestinal flora, mucosal barrier, intestinal epithelial cells, and mucosal immune system^19,20^. TcdA and TcdB have multiple effects on the innate immune system, including inducing expression of numerous pro-inflammatory mediators (e.g., cytokines, chemokines and neuroimmune peptides) and the recruitment and activation of a variety of innate immune cells^21,22^. Adaptive immunity is also sufficient to provide some protection from CDI, likely via antibody-mediated neutralization of TcdA and TcdB^23–26^. These immune markers also have the potential to act as clinically useful diagnostic markers of CDI.

Exposure to toxinogenic *C. difficile* can lead to a range of clinical outcomes ranging from asymptomatic colonization to mild diarrhea and more severe disease syndromes such as pseudomembranous colitis, toxic megacolon, bowel perforation, sepsis, and death^27,28^. Asymptomatic *C. difficile* carriage is characterized by *C. difficile* colonization in the absence of symptoms of infection. Previous studies suggest that *C. difficile* asymptomatic carriers have the potential to contribute to *C. difficile* transmission and hospital-onset CDI in inpatient facilities, as carriers can shed spores into the hospital environment^29,30^.

The diagnosis of CDI is based on clinical signs and symptoms in combination with laboratory testing. Several diagnostic laboratory tests are available including enzyme immunoassays (EIA) for TcdA and TcdB, nucleic acid amplification tests (NAAT), selective toxinogenic culture, cell cytotoxicity neutralization assay, and glutamate dehydrogenase EIA^31–33^. However, currently available approaches do not accurately differentiate CDI from diarrhea with another cause in a patient colonized with toxinogenic *C. difficile*. Over-diagnosis of disease could result in overtreatment of CDI, delayed recognition of other causes of illness, and unnecessary antibiotic exposures^34^.

Machine learning has a great impact in many areas of medical research, as it offers a principled approach for developing sophisticated, automatic, and objective algorithms for analysis of complex data. Indeed, previous studies indicate that supervised learning can be successfully employed for clinical disease assessment for diverse disorders including Parkinson’s disease^35^, diabetes^36^, inflammatory bowel disease^37^ and glaucoma^38^. In our previous work, we found that specific immune markers, particularly G-CSF, can be used to distinguish adults with CDI from other groups including asymptomatic carriers and NAAT-negative patients with and without diarrhea^39^. Here, we integrate the host immune marker data and newly obtained gut microbiome data from subjects of the same cohort to build classification models to optimally distinguish CDI from other groups. Our aim is to identify consistent biological signatures for highly accurate diagnosis of CDI.

## RESULTS

### Baseline demographic and clinical characteristics of participants

Our clinical cohort consists of 243 well-characterized recruited participants, who were divided into four groups (see Materials and Methods)^39^: (1) Control: subjects without diarrhea and with NAAT-negative stool (n=47); (2) Non-CDI Diarrhea: subjects with diarrhea but NAAT-negative stool (n=44); (3) Asymptomatic Carriage: subjects without diarrhea but with NAAT-positive stool (n=40); (4) CDI: subjects with diarrhea and NAAT-positive stool (n=112). The first three groups can be combined as the Non-CDI group. The entire clinical cohort had a mean ± SD age of 63.66 ± 14.85 year and was 48.15% female. Demographic data of the cohort are summarized in Table 1. In total, 187 participants (76.95%) had both gut microbiome and immune marker data available (see Table S1).

**Table 1.** Demographic characteristics of the enrolled subjects.

| **Characteristics** | **NAAT negative** |  | **NAAT positive** |  |
| --- | --- | --- | --- | --- |
|  | **Control**<br>**(n=47)** | **Non-CDI**<br>**Diarrhea (n=44)** | **Asymptomatic**<br>**Carriage (n=40)** | **CDI (n=112)** |
| **Sex** |  |  |  |  |
| Female | 14 (29.79%) | 22 (50.00%) | 20 (50.00%) | 61 (54.46%) |
| Male | 33 (70.21%) | 22 (50.00%) | 20 (50.00%) | 51 (45.54%) |
| **Age, Avg ± SD** | 62.40 ± 12.33 | 63.07 ± 13.15 | 62.15 ± 17.25 | 64.99 ± 15.62 |
| **Ethnicity** |  |  |  |  |
| Hispanic | 1 (2.13%) | 3 (6.82%) | 1 (2.50%) | 6 (5.36%) |
| Non-Hispanic | 38 (80.85%) | 37 (84.09%) | 31 (77.50%) | 96 (85.71%) |
| Unknown | 8 (17.02%) | 4 (9.09%) | 8 (20.00%) | 10 (8.93%) |
| **Race** |  |  |  |  |
| White | 33 (70.21%) | 28 (63.64%) | 28 (70.00%) | 89 (79.46%) |
| Other | 4 (8.51%) | 10 (22.73%) | 3 (7.50%) | 23 (20.54%) |
| Unknown | 10 (21.28%) | 6 (13.64%) | 9 (22.50%) | 0 (0.00%) |

### Microbial community structure

To compare the overall microbial community structure of the four groups, we first calculated the alpha diversity (i.e., the within-sample taxonomic diversity) of each sample at the genus level using four different measures: *taxa richness* (the observed number of different taxa present in the sample), *Chao1* (abundance-based estimator of taxa richness), *Evenness* (the uniformity of the population size of each taxa present in the sample), and *Shannon diversity index* (estimator of taxa richness and evenness: more weight on richness). (See Materials and Methods for detailed definitions of those alpha diversity measures.) As shown in Fig. 1, We found that richness indices (taxa richness and Chao1) did not differ significantly among these groups. The gut microbiota of Non-CDI Diarrhea subjects showed lower evenness than that of the Control group. Shannon diversity was significantly lower in the Non-CDI Diarrhea and CDI groups than in the Control group.

**Fig. 1.**
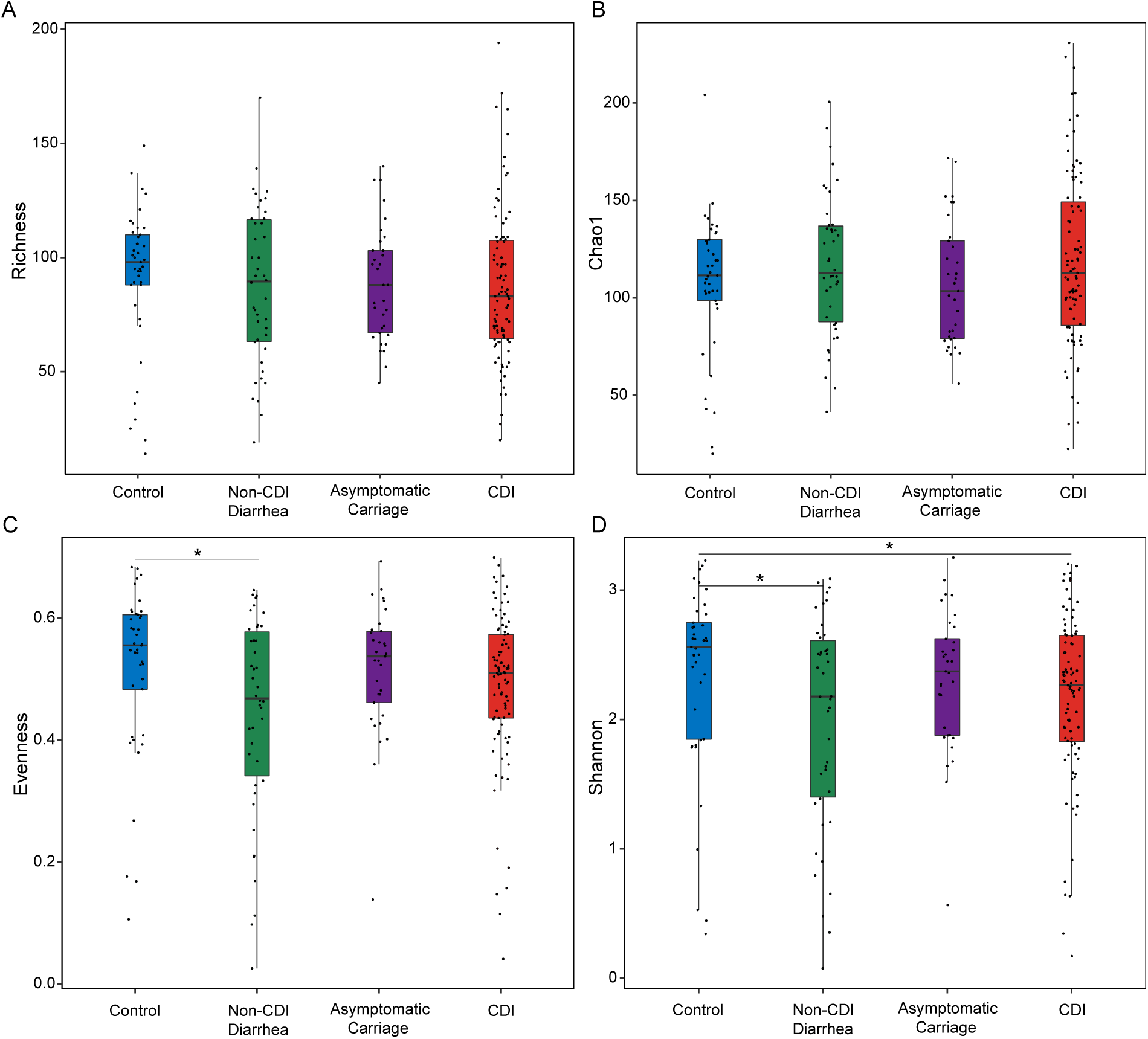
Comparing the alpha diversity of the gut microbiota of subjects with different. ***C. difficile* infection/colonization statuses (Control, Non-CDI Diarrhea, Asymptomatic Carriage, and CDI) using different alpha diversity measures**. (**A**) Taxa richness. (**B**) Chao1. (**C**) Evenness. (**D**) Shannon index. Each dot represents the alpha diversity value of a particular subject’s gut microbiota. Statistical significance was determined by Mann–Whitney test, *P<0.05.

To determine whether the gut microbial compositions of participants are affected by *C. difficile* infection/colonization status, we performed Principal Coordinates Analysis (PCoA) at the genus level using Bray-Curtis dissimilarity (which is a beta diversity measure to quantify the between-sample compositional dissimilarity). We found no distinct clusters corresponding to the four different phenotype groups, implying that the gut microbial compositions of participants from the four groups are not significantly different (Fig. 2A). Interestingly, by directly comparing the beta diversity of each group, we did find that the CDI group displays higher beta diversity than other groups (Fig. 2B), indicating that the microbial compositions of participants within the CDI group vary more prominently than other groups. Permutational multivariate analysis of variance (PERMANOVA) showed that the overall bacterial composition differed significantly among different groups based on the CDI status (*P* < 0.001; Table S2), whereas other host factors such as age, sex, race and ethnicity had no significant effect on the microbiome composition.

**Fig. 2.**
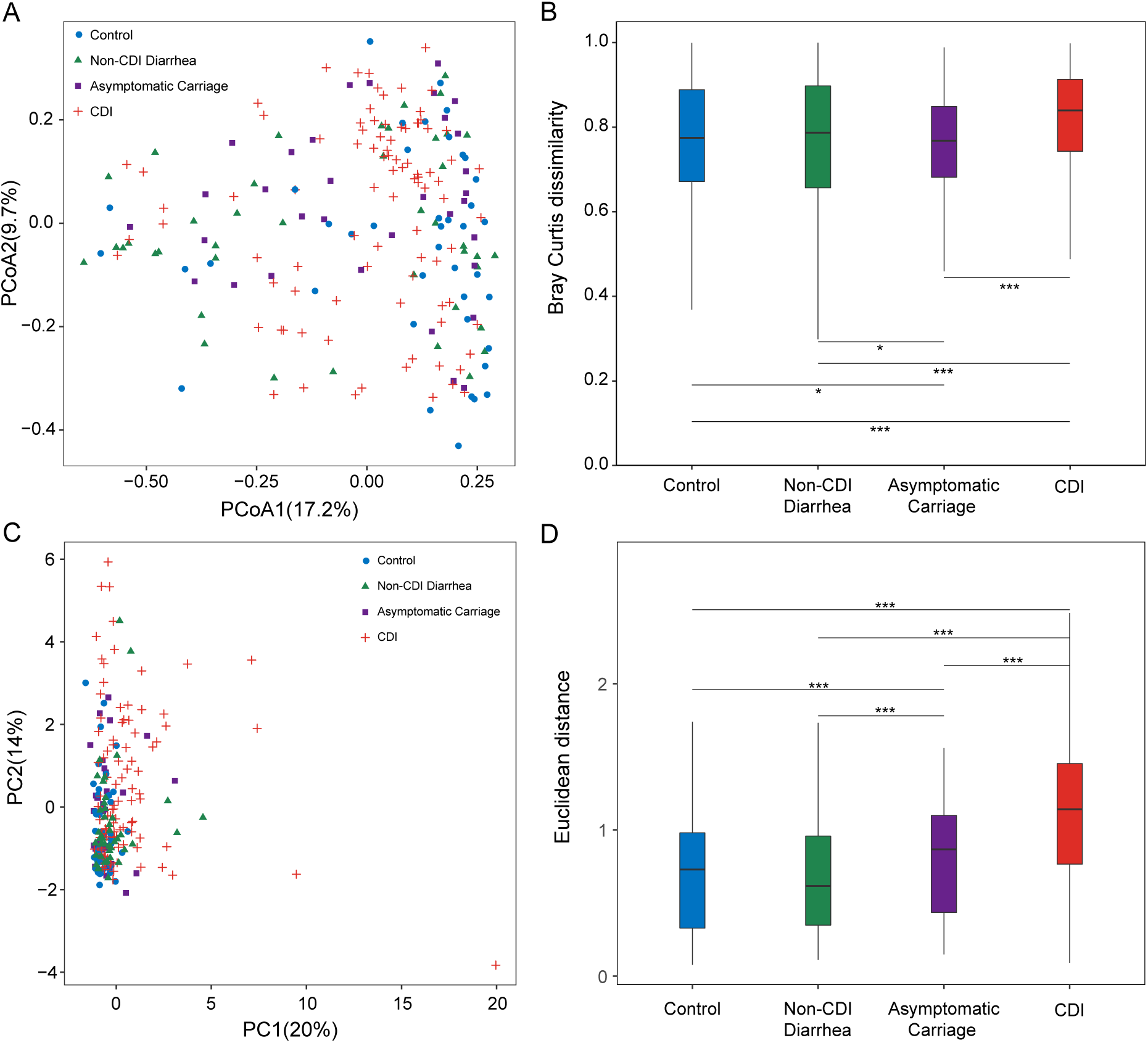
Ordination analysis and beta diversity comparison of the gut microbiota (and host immune markers) for subjects with different. ***C. difficile* infection/colonization statuses (Control, Non-CDI Diarrhea, Asymptomatic Carriage, and CDI)**. (**A**) Principal Coordinates Analysis (PCoA) plot based on Bray-Curtis dissimilarities of microbial compositions. (**B**) Boxplot of the gut microbiome Bray-Curtis dissimilarity between subjects within each group. (**C**) Principle component analysis (PCA) plot of host immune marker concentrations. (**D**) Boxplot of the Euclidean distance for the host immune markers of subjects within each group. Statistical significance was determined by Mann–Whitney test, \**P*< 0.05, \*\**P*< 0.01, \*\*\**P*<0.001.

To identify microbiome markers (i.e., certain taxa with very high discriminatory ability) to differentiate those different phenotype groups, we performed differential abundance analysis. In particular, we used ANCOM^40^ (analysis of composition of microbiomes) with a Benjamini-Hochberg correction, and adjusted for age and sex. We found that the abundances of 15 genera were significantly different between CDI and Asymptomatic Carriage groups (Fig. 3A and Table S3). Among the 15 genera, 4 of them (*Veillonella, Enterobacter, Granulicatella* and *Dialister*) of these genera were enriched in the CDI group, while the other 11 genera *(Lactococcus, Dorea, Moryella, [Ruminococcus]_gauvreauii_group, Stenotrophomonas, Agathobacter, Blautia, Sellimonas, Eggerthella, Faecalitalea* and *Lachnospiraceae UCG-008*) were enriched in the Asymptomatic Carriage group. We also found 16 differentially abundant genera between the Non-CDI Diarrhea group and the CDI group (Fig. 3B and Table S4). Of these, 10 genera (*Clostridioides, Enterobacter, Epulopiscium, Escherichia-Shigella, Eisenbergiella, Dialister, Ruminiclostridium, Fusobacterium, Klebsiella* and *Veillonella*) were enriched in the CDI group, and the other 6 genera (*[Eubacterium]_hallii_group*, *Collinsella*, *Agathobacter*, *Dorea*, *Stenotrophomonas and Streptococcus*) were enriched in the Non-CDI Diarrhea group. ANCOM analysis also enabled us to identify 40 genera (including *Clostridioides* and *Veillonella*) that have significant differential abundances between the CDI group and the whole Non-CDI group (Fig. 3C and Table S5). Note that a total of 6 differentially abundant genera were identified from all the three comparisons: CDI vs. Asymptomatic Carriage; CDI vs. Non-CDI Diarrhea; CDI vs. Non-CDI. Among them, *Veillonella, Enterobacter* and *Dialister* were enriched in the CDI group, while *Dorea, Stenotrophomonas* and *Agathobacter* were depleted in the CDI group.

**Fig. 3.**
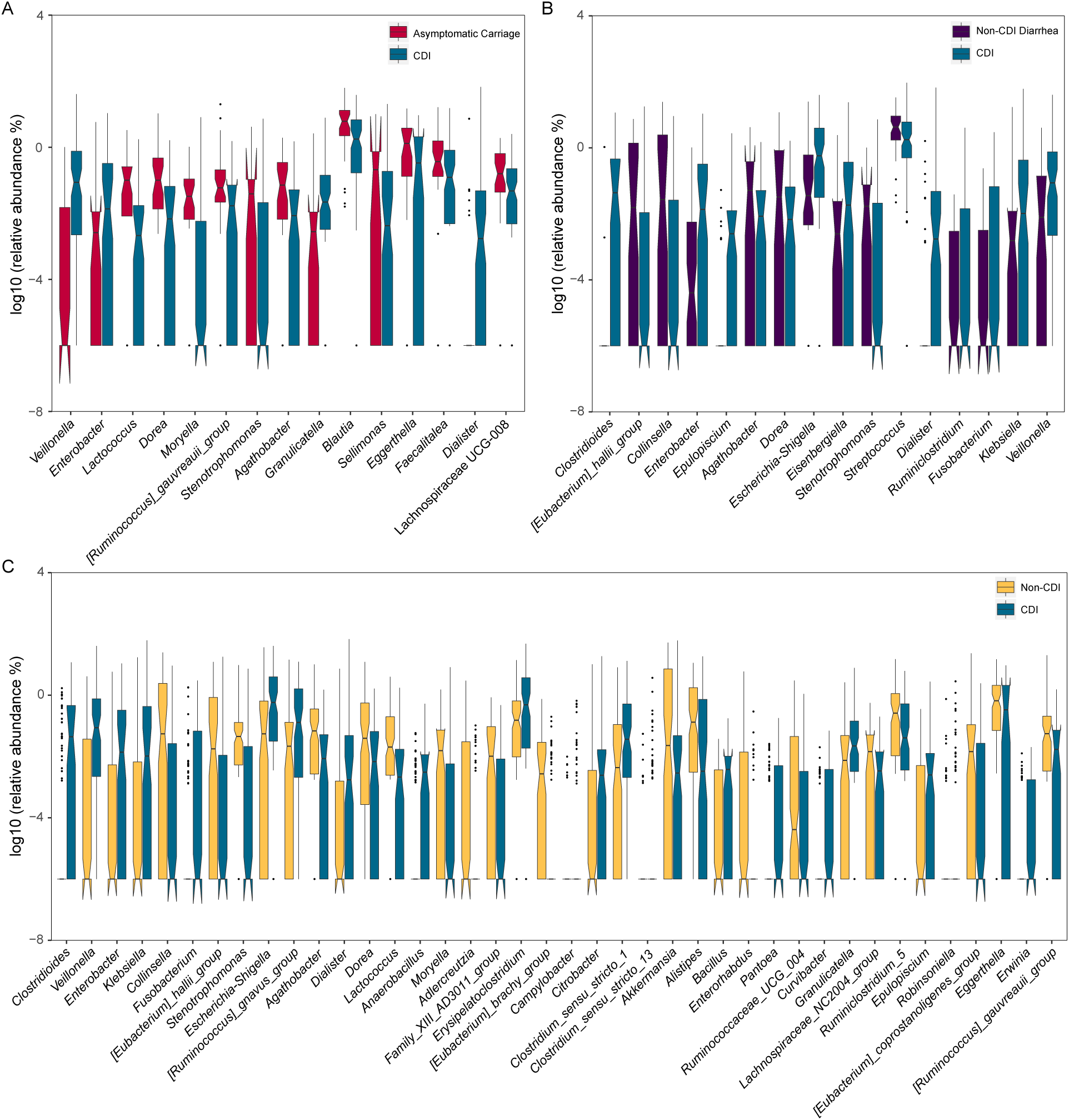
Relative abundances of differentially abundant genera identified by ANCOM in comparing different groups. (**A**) CDI vs. Asymptomatic Carriage. (**B**) CDI vs. Non-CDI Diarrhea. (**C**) CDI vs. Non-CDI. The top differentially abundant taxa were ranked based on their W statistics (from left to right). The relative abundance (%) are plotted on log10 scale. The notches in the boxplots show the 95% confidence interval around the median.

### Microbial correlation networks

To compare the microbial communities of the four groups at the network-level, we constructed the genus-level microbial correlation network for each group using SparCC^41^ (sparse correlations for compositional data). We found that the microbial correlation network of the CDI group has quite different structure compared to other groups (Fig. 4). In order to quantify the difference of the network structure, we calculated the number of nodes, number of edges, average degree (the average number of connections per node), graph density (measure of how close the network is to a complete graph), clustering coefficient (measure of how complete the neighborhood of a node is) and modularity (measure of how well a network decomposes into modular communities) (Table S6). In general, compared with the networks of other groups, the network of the CDI group has fewer nodes and edges, lower average degree, but higher modularity. These indicate that the overall microbial correlations in the CDI group are much weaker than those in other groups.

**Fig. 4.**
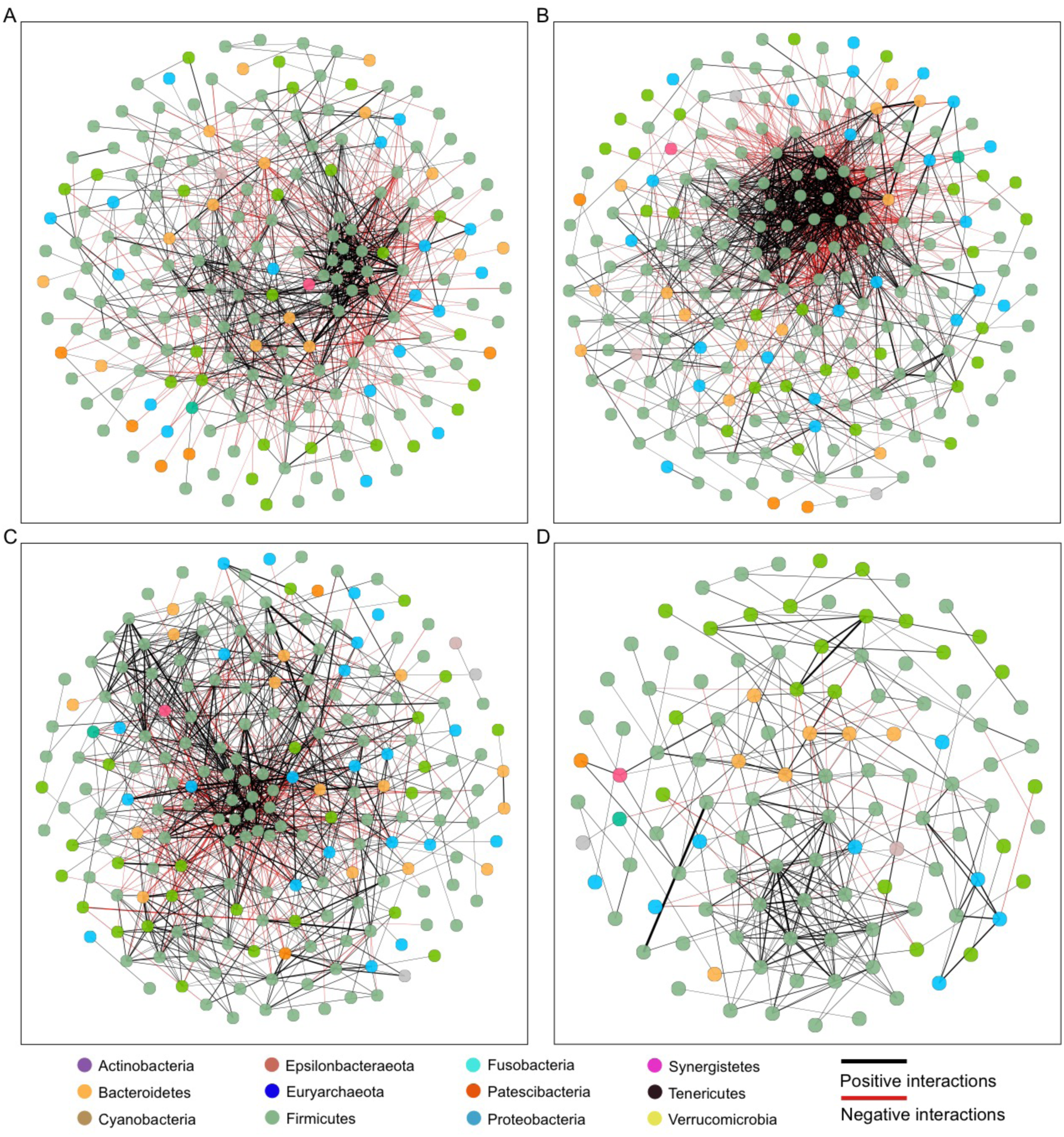
Microbial correlation networks of different groups. (**A**) Control. (**B**) Non-CDI Diarrhea. (**C**) Asymptomatic Carriage. (**D**) CDI. Nodes represent genera and are colored based on their phylum. Edges represent microbial correlations: green/red means positive/negative correlations, respectively. Edge thickness indicates correlation strength, and only the high-confidence interactions (*p*-value < 0.05) with high absolute correlation coefficients (> 0.3) were presented. For each group, we further identified the top-three most connected genera/nodes. They are *Ruminococcus_1, Roseburia* and *Lachnospiraceae_UCG-008* for the Control group, *[Ruminococcus]_torques_group, [Eubacterium]_hallii_group* and *Blautia* for the Non-CDI Diarrhea group, *Ruminiclostridium_5*, *Enterococcus* and *Lachnospiraceae_UCG_008* for the Asymptomatic Carriage group, and *Alistipes, Ruminiclostridium_5* and *Lachnoclostridium* for the CDI group.

To analyze these patterns in more detail, we used NetShift^42^ to identify potentially important “driver” taxa responsible for the change of microbial correlations. This analysis revealed 24 potential driver taxa linked with the change of microbial correlations between CDI and Asymptomatic Carriage groups (Fig. S1). The top driver taxa were *Alistipes, Clostridioides, Desulfovibrio, Eggerthella, Erysipelatoclostridium, Klebsiella, Odoribacter Proteus, [Ruminococcus]_torques_group, Streptococcus, Vagococcus* and *Veillonella*. We then identified 24 genera as potential driver taxa underlying the change of microbial correlations between CDI and Non-CDI Diarrhea groups (Fig. S2). The top driver taxa were *Alistipes, Buttiauxella, Citrobacter, Clostridium_sensu_stricto_13*, *Desulfovibrio, Klebsiella, Oscillibacter, Phascolarctobacterium*, *Streptococcus* and *Veillonella*. Finally, Netshift analysis revealed 38 potential driver taxa underlying the change of microbial correlations between CDI and Non-CDI groups. The top driver taxa were *Bifidobacterium, Clostridioides, Klebsiella, Oscillibacter, Streptococcus* and *Veillonella* (Fig. S3). Together, these results suggested that certain bacterial taxa (e.g., *Clostridioides, Klebsiella, Streptococcus* and *Veillonella*) could play an important role in driving the changes of microbial correlations in subjects with different *C. difficile* infection/colonization status.

### Host immune markers and CDI

To determine the systemic levels of proinflammatory cytokines in CDI, we measured the circulating levels of granulocyte-colony stimulating factor (G-CSF), interleukin-1β (IL-1β), IL-2, IL-4, IL-6, IL-8, IL-10, IL-13, IL-15, monocyte chemoattractant protein-1 (MCP-1), vascular endothelial growth factor-A (VEGF-A), and tumor necrosis factor-alpha (TNF-α) as previously reported^39^. Serum concentrations of immunoglobulin A (IgA), IgG, and IgM antibodies against *C. difficile* toxin A and toxin B were measured by semi-quantitative enzyme-linked immunosorbent assay (ELISA). We previously demonstrated specific markers of these innate and adaptive immunity that can distinguish CDI from each of the other three groups^39^. In the current study, we are particularly interested in comparing the CDI group and the combined Non-CDI group. Based on the Mann-Whitney U test, we identified in total 11 immune markers that displayed significantly different concentrations in these two groups, including G-CSF, IL-4, IL-6, IL-8, IL-10, IL-15, TNF-α, MCP1, IgA anti-toxin A and B, and IgG anti-toxin A in blood (Table S7). All of these immune markers had higher concentrations in the CDI group than in the Non-CDI group. Host immune marker variations between samples were evaluated using the Principal Component Analysis (PCA) (Fig. 2C). PCA plot showed no clear clustering of those subjects based on immune marker concentrations. However, boxplot of Euclidean distance of immune marker profiles from CDI patients showed higher within-group variation than that in all the other three groups (Fig. 2D). PERMANOVA analysis indicated that the immune homeostasis was significantly different among different groups based on the CDI status (P = 0.016; Table S2). But age, gender, race and ethnicity did not have significant effects on the host immune marker levels.

### Interplay between gut microbiome and host immune markers

To reveal the interplay between the gut microbiome and the host immune system, we calculated the correlations between microbial compositions (at the genus level) and the circulating levels of host immune markers for each of the four groups (Spearman correlation with Benjamini-Hochberg correction). The results are shown in Fig. 5 and Fig. S4. For the Control group, the most significant associations were identified as *Chiristensenellaceae R-7 group* negatively associated with TNFα, *Bifidobacterium* positively associated with VEGFA and IL-13, *Rothia* positively associated with IL-15, and *Veillonella* positively related with IL-4 (Fig.5A and Fig. S4). For the Non-CDI Diarrhea group, *Ruminococcaceae UCG-011* was negatively correlated with IL-8 and IL-6, *Defluviitaleaceae UCG-011* was positively correlated with IL-1b, and *Blautia* was negatively correlated with MCP1 levels (Fig. 5B). For the Asymptomatic Carriage group, we found that *Lactobacillus* was negatively associated with VEGFA, *Akkermansia* was positively associated with IL-6, and *Enterococcus* was positively related to TNFa (Fig. 5C). For the CDI group, negative associations involved *Akkermansia* and IL-10, *Lactococcus* and G-CSF, while positive associations involved *Lactobacillus* and IgG and IgA anti-toxin B (Fig. 5D). Interestingly, none of these most significant associations was universally present across different groups. This indicated that the interactions between gut microbiota and host immunological markers can be very sensitive to the status of *C. difficile* colonization and infection. More importantly, this result implies that the integration of gut microbiota and host immune markers might be quite useful for highly accurate diagnosis of CDI.

**Fig. 5.**
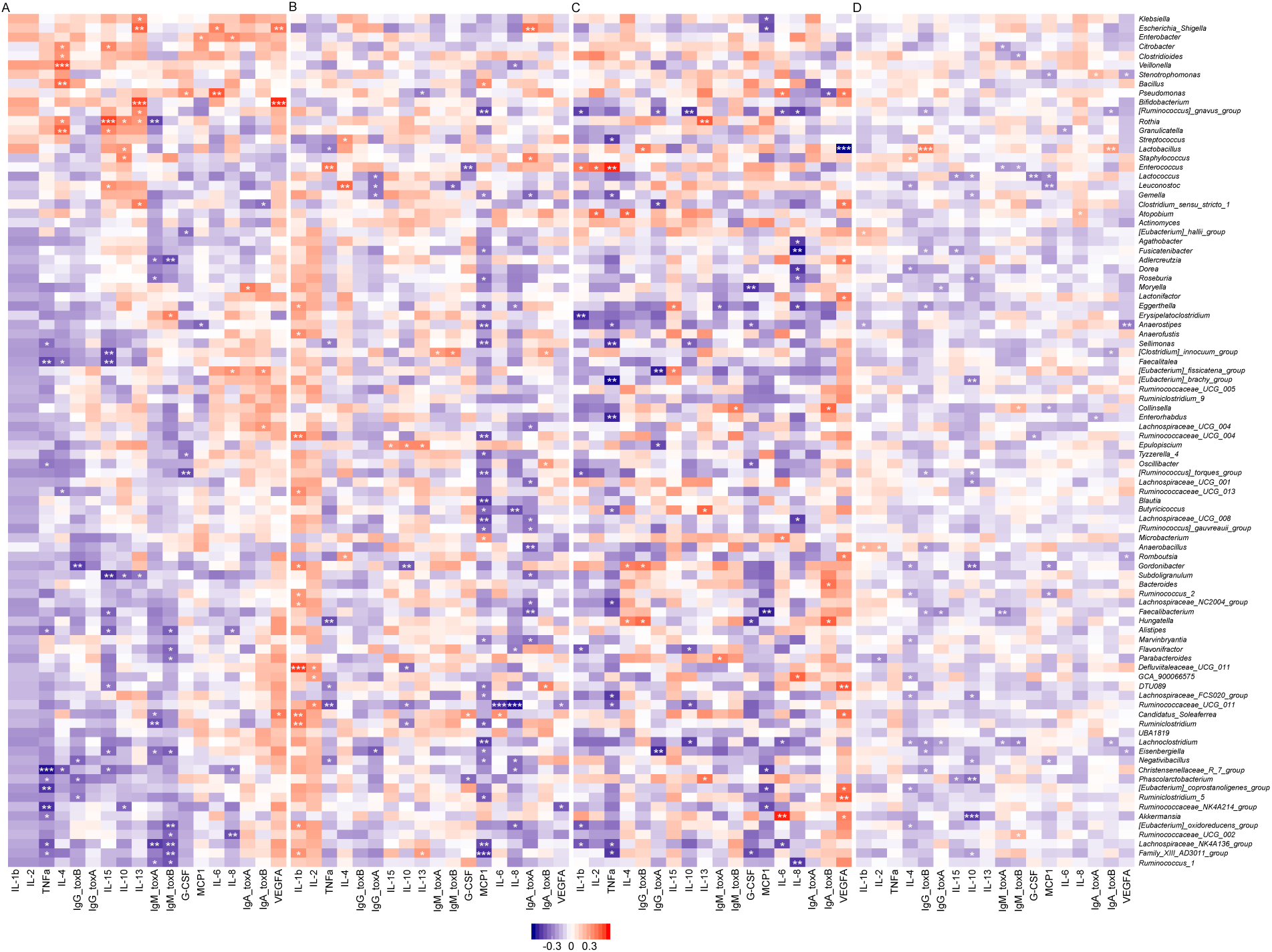
Correlations between gut microbial abundances and host immune markers in different groups, quantified by Spearman correlation with Benjamini-Hochberg correction. (**A**) Control. (**B**) Non-CDI Diarrhea. (**C**) Asymptomatic Carriage. (**D**) CDI. Rows represent genera; columns represent immune markers. The layout of the heatmap is followed the hierarchical clustering results of Control cohort (see Fig.S4). Red/blue represents positive/negative correlation, respectively. The intensity of the colors denotes the strength of the correlation. *α<0.05, **α<0.01, ***α< 0.001.

### Diagnostic accuracy for CDI classification based on host immune markers and gut microbiota

To determine whether host immune markers or gut microbiota could serve as biomarkers to classify subjects into different groups, we constructed a multi-class classifier based on random forests (RF). One of the most popular performance metrics of a classifier is the Area Under the receiver operating characteristic Curve (AUC). The performance of a multi-class classifier is measured by both micro-average and macro-average AUCs. (For micro-average AUC, we calculated the AUC from the individual true positive rates and false positive rates of the multi-class model. For the macro-average AUC, we calculated the AUC independently for each class and then took the average.) We considered three different feature types: (1) host immune maker concentrations alone; (2) gut microbial compositions alone; and (3) the integration of (1) and (2) in our classification analysis. To eliminate confounding effects, we excluded the genus *Clostridioides* from our classification analysis. The immune marker-based classifier achieved macro-average AUC ~ 0.827 and micro-average AUC ~ 0.828 (Fig. S5A), which are quite comparable to the performance of microbiota-based classifier (Fig. S5B). Interestingly, integrating immune marker with gut microbiota showed much better classification performance (macro-average AUC ~ 0.926 and micro-average AUC ~ 0.869) (Fig. S5C).

We further performed binary classifications to distinguish CDI subjects from Asymptomatic Carriage, Non-CDI Diarrhea, and Non-CDI subjects, using different feature types (Fig. 6). The goal of this analysis was to assess whether any single taxon or immune marker could reliably differentiate CDI status. The importance of each feature was quantified by the Mean Decrease in Accuracy (MDA) of the classifier due to the exclusion (or permutation) of this feature. The more the accuracy of the classifier decreases due to the exclusion (or permutation) of a single feature, the more important that feature is deemed for classification of the data.

**Fig. 6.**
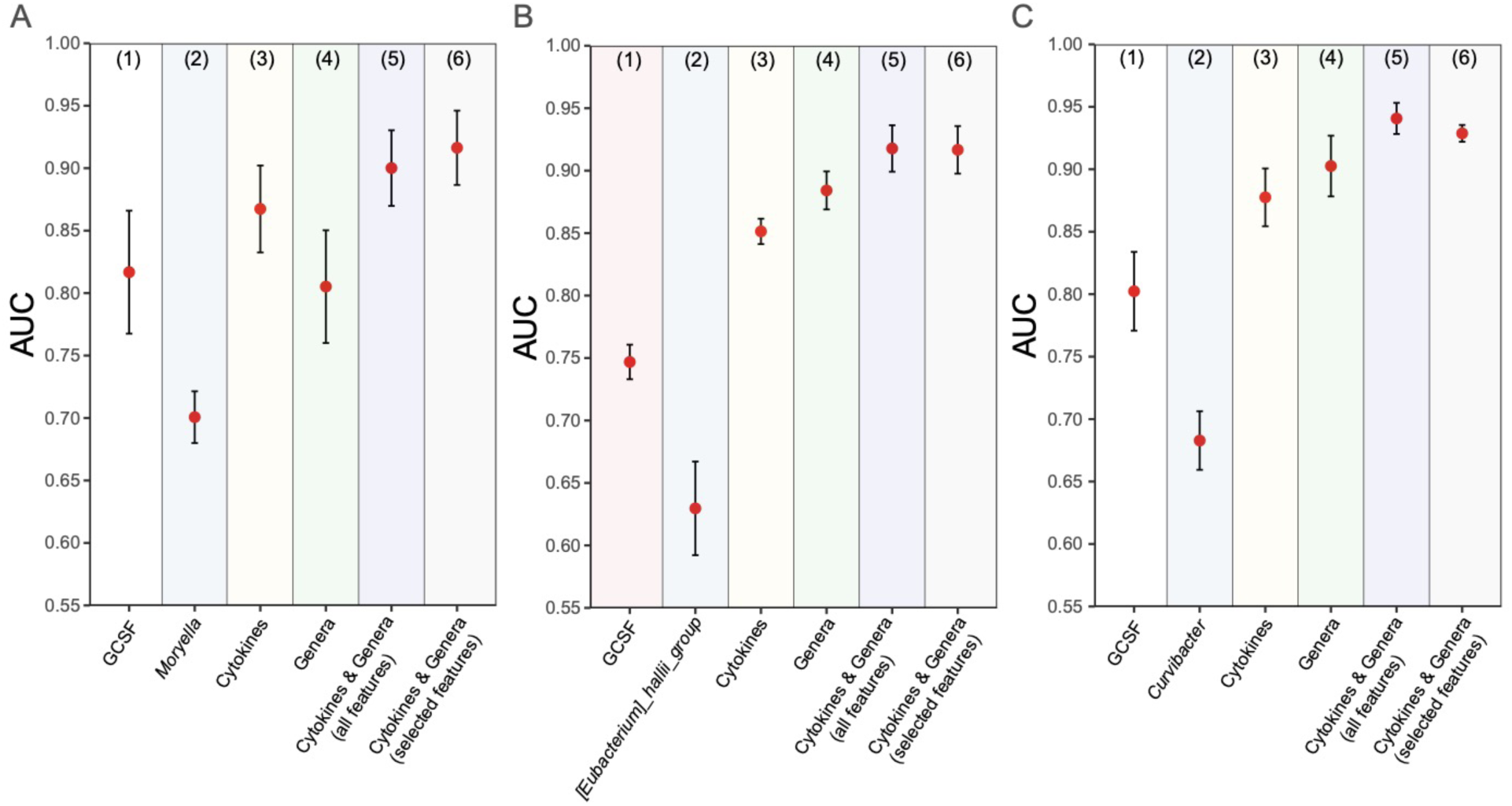
The performance of RF-based classification models based on various types of features in differentiating CDI from other groups. (**A**) CDI vs. Asymptomatic Carriage. (**B**) CDI vs. Non-CDI Diarrhea. (**C**) CDI vs. Non-CDI. For each classification task, we used different types of features: (1) the top-1 immune marker feature (based on mean decrease accuracy); (2) the top-1 genus feature; (3) all immune markers; (4) all genera; (5) integration of all immune markers and genera; (6) selected features from the set of all immune markers and genera. Error bars represent the standard errors of the means (SEM).

In the classification of CDI vs. Asymptomatic Carriage, we found that G-CSF and *Moryella* were the most important immune and microbial features, respectively (Fig. S6:A-B). But the classification based on G-CSF (or *Moryella*) alone did not yield very high performance: mean AUC ~ 0.817 (or 0.701), respectively (Fig. 6:A1-A2). When we used all the immune markers (or all the genera) as features, we achieved mean AUC ~ 0.867 (or 0.805), respectively (Fig. 6:A3-A4). Interestingly, when we integrated all the host immune markers and gut microbial composition data together, we achieved a much higher performance with mean AUC ~ 0.900 (Fig. 6:A5). In order to select a subset of features that is as discriminatory as the whole set of features, we followed the “1-SE” rule (i.e., one chooses the model with fewest features such that its classification performance is less than one standard error away from that of the model with all the features), and selected the following 4 features: 2 bacterial genera (*Moryella* and *Veillonella*) and 2 immune markers (G-CSF and IL-6) in classifying CDI and Asymptomatic Carriage groups (Fig. S6:G-J). The RF classifier with those selected features displayed an outstanding classification performance, with mean AUC ~ 0.916 (Fig. 6:A6). Note that a significant negative correlation between *Moryella* and G-CSF was found in the Asymptomatic Carriage group (Fig. 5C), which might contribute to the outstanding performance of the RF classifier with *Moryella* and G-CSF as selected features.

In the classification of CDI vs. Non-CDI Diarrhea groups, we found that G-CSF and *[Eubacterium]_hallii_group* are the top immune and microbial features, respectively (Fig. S6:C-D). But the classification based on G-CSF (or *[Eubacterium]_hallii_group*) alone did not perform very well: mean AUC ~ 0.747 (or ~ 0.630), respectively (Fig. 6:B1-B2). When we used all the immune marker (or all the microbial genera) as features, we achieved mean AUC ~ 0.851 (or ~ 0.884), respectively (Fig. 6:B3-B4). By integrating all features from both host immune marker and gut microbial genera, we further improved the classification performance to mean AUC ~ 0.918 (Fig. 6:B5). Following the “1-SE” rule, we selected the following 5 features: 3 genera: *Enterococcus, Epulopiscium and [Eubacterium]_hallii_group*; and 2 immune markers: G-CSF and IgA anti-toxin A (Fig. S6:H-K). The RF classifier with those selected features achieved mean AUC ~ 0.917 (Fig. 6:B6), which is quite comparable to that of using all the features. Note that *Enterococcus* was found to be significantly associated with G-CSF in the Non-CDI Diarrhea group (Fig. 5B). This might partially explain the outstanding performance of the RF classifier with *Enterococcus* and G-CSF as selected features.

In the classification of CDI vs. Non-CDI groups, we found that G-CSF and *Curvibacter* are the top immune and microbial features, respectively (Fig. S6:E-F). Classification based on G-CSF (or *Curvibacter*) alone achieved mean AUC ~ 0.802 (or ~ 0.683), respectively (Fig. 6:C1-C2). When we used all the immune marker (or all the microbial genera) as features, we achieved mean AUC ~ 0.878 (or ~ 0.903), respectively (Fig. 6:C3-C4). Integrating all features from both host immune marker and gut microbial genera, we further improved the classification performance to mean AUC ~ 0.941 (Fig. 6:C5). Following the “1-SE” rule, we selected the following 10 features: 6 genera: *Stenotrophomonas, Curvibacter, Enterobacter*, *Anaerobacillus, Fusobacterium* and *Veillonella*; and 4 immune markers: G-CSF, IL-6, TNF-α and IgA anti-toxin B (Fig. S6:I-L). Classification with those well selected features achieved mean AUC ~ 0.929 (Fig. 6:C6).

### Using symbolic classification to derive diagnostic scores

The outstanding classification results based on well-selected features prompt us to derive simple mathematical models for CDI diagnosis. To achieve that, we leveraged symbolic classification (SC)^43,44^, a genetic programming technique that automatically searches the space of mathematical expressions to find the model that best fits a given dataset. The fitness function in SC is a maximization function, and the number of generations is chosen based on the saturation of the fitness score (Fig. S7). Using the same set of selected features and trained with the entire dataset, the SC model outperformed logistic regression (LR) in differentiating CDI from Asymptomatic Carriage (or Non-CDI Diarrhea, or Non-CDI), based on various performance metrics: Accuracy, Precision, Recall and F1-score (see Table 2).

**Table 2.** Diagnostic scores derived from symbolic classification (SC) and logistic regression (LR). For each subject *i*, we calculate his/her diagnostic score *f*(*i*) (or *p*(*i*)) based on one of the following formulas derived from SC (or LR), respectively. For SC, the class of subject *i* is CDI if *f*(*i*) *>* 0; or Asymptomatic Carriage (or Non-CDI Diarrhea, Non-CDI) if *f*(*i*) ≤ 0. For LR, the class of subject *i* is CDI if *p*(*i*) *≥* 0.5; or Asymptomatic Carriage (or Non-CDI Diarrhea, Non-CDI) if *p*(*i*) *<* 0.5. Here, both *f*(*i*) and *p*(*i*) were learned from the entire dataset. Features used here include: *x*_1_: GCSF; *x*_2_: IgA_toxA; *x*_3_: IgA_toxB; *x*_4_: IL6; *x*_5_: TNFα; *x*_6_: *Anaerobacillus*; *x*_7_: *Curvibacter*; *x*_8_: *Enterobacter*; *x*_9_: *Enterococcus*; *x*_10_: *Epulopiscium*; *x*_11_: *[Eubacterium]_haillii_group; x*_12_: *Fusobacterium*; *x*_13_: *Moryella*; *x*_14_: *Stenotrophomonas; x*_15_: *Veillonella*. In particular, for each classification task (regardless of using SC or LR), the following selected features were: (1) CDI vs. Asymptomatic Carriage: *x*_1_, *x*_4_, *x*_13_ and *x*_15_; (2) CDI vs. Non-CDI Diarrhea: *x*_1_, *x*_2_, *x*_9_, *x*_10_, and *x*_11_; (3) CDI vs. Non-CDI: *x*_1_, *x*_3_, *x*_4_, *x*_5_, *x*_6_, *x*_7_, *x*_8_, *x*_12_, *x*_14_ and *x*_15_. Note that in the calculation of precision, recall and Fl-score, we can treat either CDI (or Asymptomatic Carriage, Non-CDI Diarrhea, Non-CDI) as the true positive. Results shown in the parenthesis represent the latter case.

| Model | Diagnostic | Formula | Accuracy | Precision | Recall | F1-score |
| --- | --- | --- | --- | --- | --- | --- |
| SC | CDI vs. Asymptomatic Carriage | $f(i) = x_1 * x_{15}(x_1^3 - 0.2 * x_{13} + 0.4) + 1.1 * x_1 - 0.1 * x_4 - 18.25$ | 0.896 | 0.914<br>(0.840) | 0.949<br>(0.75) | 0.931<br>(0.792) |
| | CDI vs. Non-CDI Diarrhea | $f(i) = x_9 * x_2(0.5 * x_{10} - 1) + x_{11}(0.02 * x_{11} - x_1) + x_2 \left(1 - \frac{10}{x_1}\right) - \frac{0.003}{x_9}$ | 0.900 | 0.946<br>(0.826) | 0.897<br>(0.905) | 0.921<br>(0.864) |
| | CDI vs. Non-CDI | $f(i) = x_1 * x_3(0.2 * x_1 * x_5 * x_6 * x_{14} + 0.04 * x_1 * x_7 + 0.3 * x_1 * x_{15} * x_8^4 + x_1 * x_{12}(0.5 * x_7 + x_1 * x_{14}) + x_7(0.1 * x_4 - x_6) + x_{14}(x_{14} - 2)$ | 0.882 | 0.889<br>(0.878) | 0.821<br>(0.927) | 0.853<br>(0.902) |
| LR | CDI vs. Asymptomatic Carriage | $\log\left(\frac{p(i)}{1-p(i)}\right) = 0.66725 - 0.04442 * x_1 + 0.01022 * x_4 + 7.51484 * x_{13} - 85.00213 * x_{15}$ | 0.830 | 0.895<br>(0.667) | 0.872<br>(0.714) | 0.883<br>(0.690) |
| | CDI vs. Non-CDI Diarrhea | $\log\left(\frac{p(i)}{1-p(i)}\right) = 0.01974 - 0.002084 * x_1 - 0.02391 * x_2 + 1.895 * x_9 - 12740 * x_{10} + 163.9 * x_{11}$ | 0.800 | 0.814<br>(0.765) | 0.897<br>(0.619) | 0.854<br>(0.684) |
| | CDI vs. Non-CDI | $\log\left(\frac{p(i)}{1-p(i)}\right) = 2.122 - 0.01002 * x_1 + 0.01833 * x_3 - 0.006334 * x_4 - 0.009566 * x_5 - 4609 * x_6 - 8576 * x_7 - 40.75 * x_8 - 101.1 * x_{12} + 32.84 * x_{14} - 43.4 * x_{15}$ | 0.813 | 0.841<br>(0.798) | 0.679<br>(0.908) | 0.752<br>(0.850) |

Indeed, as shown in Table 2, we derived a simple SC model with selected features, reaching a very high accuracy (0.896) in distinguishing CDI subjects from Asymptomatic Carriage. Basically, for each subject *i*, we calculate the diagnostic score *ƒ*(*i*) that will be used for CDI diagnosis: the class of subject *i* is CDI if *ƒ*(*i*) > 0; Asymptomatic Carriage, if *ƒ*(*i*) ≤ 0. Here,

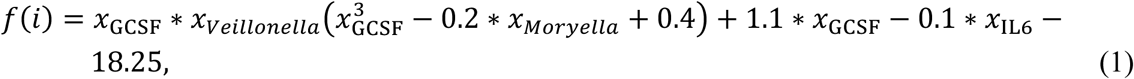

with *x_a_* representing the abundance or concentration of feature-*a* in subject-*i*. Similarly, we derived a SC model with accuracy of 0.900 in distinguishing CDI (if *ƒ*(*i*) > 0) from Non-CDI Diarrhea (if *ƒ*(*i*) < 0) with the diagnostic score

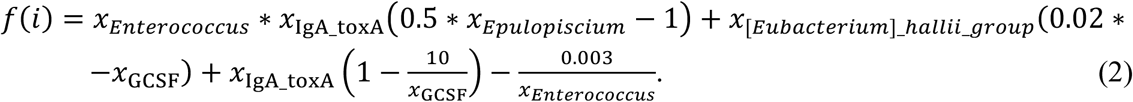

Finally, we derived a SC model with accuracy of 0.882 in distinguishing CDI (if *ƒ*(*i*) > 0) from Non-CDI (if *ƒ*(*i*) ≤ 0) with the diagnostic score

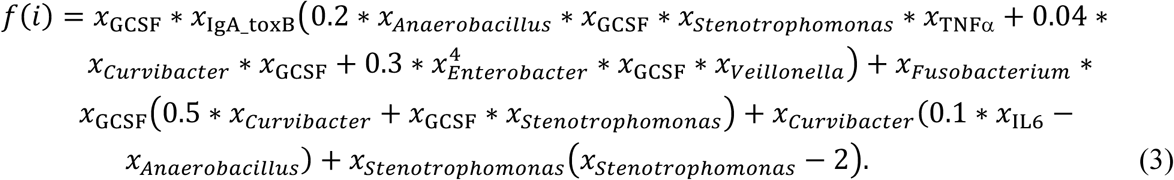

To ensure the SC models learned from the entire dataset are not overfitting, we performed cross-validation by randomly splitting the dataset to form a training set (80% of the data) and a held-out test set (20% of the data) in 10 different ways. Each time, for each classification task, we learned the SC model from the training dataset and evaluated it on the test dataset. Due to the different training sets, SC will derive different mathematical formulas (i.e., diagnostic scores). However, those SC models learned from different training datasets demonstrated quite robust performance in terms of Accuracy, Precision, Recall and F1-score (see Table S8). More importantly, even trained with less data, the SC models still outperformed LR models learned from the entire dataset.

These SC models consisted of explicit mathematical equations, which are more transparent than black-box classifiers such as RF. At the same time, the SC models are also more accurate than traditional classifiers (such as LR). The transparency and high accuracy highlight the importance of SC models in the clinical diagnosis of CDI.

## DISCUSSION

Current methods for CDI diagnosis are unable to combine high sensitivity and high clinical specificity, which can result in either underdiagnosis or overdiagnosis of CDI^45^. A more accurate diagnostic approach for CDI could optimize therapeutic decision-making and reduce transmission. Here, we employed 16S rRNA gene sequencing to profile the gut microbial compositions and combined the gut microbiome data with data from a broad panel of innate and adaptive host immune response markers to investigate the potential roles of these markers in the diagnosis of CDI. We demonstrated that the combination of host immune markers and gut microbial data can provide a potential route to optimize CDI diagnosis. Importantly, this work derived specific diagnostic models (in terms of mathematic equations) that yielded robust accuracy in differentiating CDI subjects from Asymptomatic Carriage, Non-CDI Diarrhea and Non-CDI groups.

Taxonomic diversity is a fundamental property of ecological systems. It is generally believed to be an important determinant of the structure and functioning of ecological communities^46,47^. Diversity indices have been routinely calculated in the study of human microbiome^48^. Consistent with previous studies^49–52^, we found that the gut microbiomes of CDI patients were characterized by lower Shannon diversity than that of the Control group. Interestingly, we observed an increased variation of both immune markers and gut microbial compositions in the CDI group with respective to other studied groups. This suggests that CDI is characterized by a significantly less stable microbiome and immune homeostasis. Our findings are in line with the Anna Karenina principle, which suggests that CDI linked changes in the microbiome and immune homeostasis are likely stochastic, leading to community instability^53–55^.

We were able to identify several candidate driver taxa (e.g., *Desulfovibrio, Klebsiella, Streptococcus* and *Veillonella*) that played a key role in driving the changes of microbial correlation networks between CDI and Asymptomatic Carriage (or Non-CDI Diarrhea, Non-CDI) groups. Among those driver taxa, *Desulfovibrio* has previously been shown to have a pathogenic role in ulcerative colitis due to its ability to generate sulfides^56^. *Streptococcus* has previously been shown to produce lactate thus impacting *C. difficile* TcdA production and *tcdA* expression to alleviate CDI^57^. *Klebsiella* is a Gram-negative bacterium that cause different types of healthcare-associated infections including pneumonia, bloodstream infections, and meningitis^58^. *Klebsiella* bacteria have been increasingly shown to develop antimicrobial resistance, most recently to the class of antibiotics known as carbapenems^59,60^. It is thus possible that the CDI pathogenesis is further enforced by the enrichment of antagonistic bacteria present in the gut microbiome of CDI subjects. In addition, our analysis demonstrated that the associations between host immunological markers and gut microbial compositions in the CDI group were dramatically different from those in other groups. However, further investigations are needed to determine whether these alterations are integral to the CDI pathogenesis.

The diagnosis of CDI remains challenging, especially the ability to distinguish CDI and *C. difficile* colonization^61–63^. To address this issue, we developed classification models aimed at differentiating CDI status based on host immune markers and gut microbiome data. We excluded the genus *Clostridioides* in further classification analysis to eliminate confounding effects. Evaluating the classification performance of host immune markers or/and microbiome data in multi-class models, it appears that a combination of host immune markers and gut microbiome data can further improve the accuracy of classification. More specifically, we were able to identify specific immune and microbial features that could accurately distinguish CDI subjects from Asymptomatic Carriage, Non-CDI Diarrhea, and Non-CDI subjects. In addition, most of the selected features identified by feature selection were also differentially abundant genera and differentially expressed immune markers.

From the classification of CDI and Asymptomatic Carriage, we were able to select a few features with outstanding discriminability, including *Veillonella* and *Moryella*. Interestingly, a positive relationship between *Veillonella* and CDI has been identified in recent studies^64–67^. An important role for *Veillonella* in CDI is supported by the fact that *Veillonella* species were associated with low coprostanol levels that correlated strongly with CDI^64^. A similar negative relationship between *Moryella* species and CDI has previously been observed^68^. *Enterococcus*, a feature selected from the classification of CDI vs. Non-CDI Diarrhea, has been reported to be associated with CDI due to vancomycin resistance^69^. Consistent with the findings from previous reports^70,71^, *Epulopiscium* was significantly enriched in the CDI group and played an important role in differentiating this comparison. Among those features selected from the classification of CDI and Non-CDI groups, *Enterobacter* and *Fusobacterium* have been considered as opportunistic pathogens involved in multiple diseases^72,73^.

Machine learning method has the potential to identify biomarkers and aid in the diagnosis of many diseases. However, the learnt relationships between predictors and outcome are typically non-transparent, especially non-linear methods (i.e., decision tree learning). Previous study has shown that an interpretable trees framework can extract, measure, prune, select, and summarize rules from a tree ensemble, and calculates frequent variable interactions^74^. However, these rules from tree ensembles are still too complicated to be clinically meaningful. Classical logistic regression, is one of the most common machine learning models in medicine^75^. The main drawback of LR is its failure to solve non-linear problems and it underperforms where there are multiple or non-linear decision boundaries^76^. Furthermore, the log odds scale in LR is hard to interpret^77^. Symbolic classification based on genetic programming is an automated technique to derive formulas from features for classification purpose^78^. Using the selected integrated features from the random forests model, we demonstrated that the mathematical formulas automatically derived from symbolic classification have robust diagnostic accuracy to differentiate CDI patients from Asymptomatic Carriage (or Non-CDI Diarrhea, and Non-CDI groups). Specifically, symbolic classification provides explicit mathematic formulas as its output, which significantly improves the transparency of the learned relationship between predictors and outcomes. These results hold translational promise in clinical diagnosis of CDI. Further external validation of the derived formulas will require a different cohort with the same inclusion criteria as ours. This is beyond the scope of the current work.

We previously demonstrated the potential clinical utility of a specific immunological biomarker (G-CSF) for CDI diagnosis^39^. This study leverages the newly obtained gut microbiome data from the same unique and well-characterized study cohort, allowing us to study integrated host immune marker and gut microbiome signatures. The fundamental differences between this study and our previous one are the clinical utilization of integrated immune and microbiome signatures to distinguish CDI patients from Asymptomatic Carriage, Non-CDI Diarrhea, and Non-CDI groups, and to derive diagnostic scores for CDI diagnosis. We believe that this study sets the stage to explore the potential role of an immune and microbiome-based test for CDI diagnosis. Of course, observed associations and selected features do not offer any causal relationships. Prospective studies are needed to validate the mechanism underlying the relationship between these selected features/biomarkers and the CDI infection/colonization status. The 16S rRNA sequencing may not have captured additional insights associated with the disease status available at the species or strain level. Further studies are needed to validate the clinical utility of the proposed biomarkers by metagenomics sequencing as well as metatranscriptomics, metaproteomics and metabolomics.

In summary, leveraging a well-characterized clinical cohort, we provided strong evidence that integrating gut microbiome and host immune signatures can significantly improve the CDI diagnosis. In particular, these results demonstrate that knowledge of gut microbial compositions in combination with host immune markers is beneficial in generating clinically relevant machine learning models for disease diagnosis. Indeed, the machine learning models show high diagnostic accuracy in differentiating true CDI from asymptomatic carriage of *C. difficile* and from non-*C. difficile* diarrhea, which are areas where current laboratory testing for CDI lacks adequate clinical specificity.

## MATERIALS AND METHODS

### Study cohort

The background and design of this cohort has been described in details previously^62^. Concisely, we have four groups associated with different *C. difficile* infection/colonization statuses: (1) Control: subjects without diarrhea who had screened as eligible for the asymptomatic carriage group (see below) but were NAAT-negative on research stool testing; (2) Non-CDI Diarrhea: subjects with diarrhea but NAAT-negative stool on clinical testing; (3) Asymptomatic Carriage: subjects were admitted for at least 72 hours, had received at least one dose of an antibiotic within the past 7 days, did not have diarrhea in the 48 hours prior to stool sample collection, had positive NAAT results on research stool testing and were not treated for CDI; (4) CDI: inpatients with positive clinical stool NAAT result, diarrhea, and a decision to treat for CDI. All subjects were adults (age ≥ 18 years old). Clinical serum samples were collected as discards within 24 hours of stool sample collection. In our previous study^39^, the four groups were named as (1) “no Diarrhea NAAT-Negative” = Control; (2) diarrhea NAAT-negative = Non-CDI Diarrhea; (3) Carrier-NAAT = Asymptomatic Carriage and (4) CDI-NAAT = CDI. In this work, for simplicity we used the simpler and more clearly descriptive titles.

### Serum immune marker measurement

The measurement of host serum cytokines concentrations of IL-2, IL-4, IL-6, IL-8, IL-10, IL-13, IL-15, IL-1β, G-CSF, IL-1β, MCP-1, VEGF-A, and TNF-α was performed using a Milliplex magnetic bead kit and Luminex analyzer (MAGPIX) (Millipore Sigma, Inc., Burlington, MA) as per the manufacturer’s instructions. Purified toxin A and B were separately prepared from *C. difficile* strain VPI 10463 (American Type Culture Collection 43255-FZ, Manassas, VA). Serum antibody (IgA, IgG, and IgM) levels against *C. difficile* toxins A and B were measured by semi-quantitative enzyme-linked immunosorbent assay (ELISA). All the experimental details have been reported previously^39,62^.

### Fecal DNA extraction and bacterial 16S rRNA sequencing data analysis

Stool DNA was extracted using the DNeasy PowerSoil Pro Kit (Qiagen, cat# 12888–100) in a QiaCube automated DNA extraction system (Qiagen) according to instructions. Briefly, 250mg stool was transferred into a PowerBead Pro Tube provided with the kit and 200 ug RNaseA and 800 μl of CD1 solution were added. Tubes were vortexed briefly, transferred into an adapter, and then vortexed at maximum speed for 10 min. Tubes were centrifuged at 15,000 x g for 1 min and about 500–600 μl supernatant was used for DNA extraction according to instructions. DNA were eluted in 70 μl elution solution C6 and stored at −80^0^C until use. 16S rRNA microbiome characterization was performed by sequencing the V4 region of the 16S rRNA gene using the Illumina MiSeq.^79^ Each sample was amplified using a barcoded primer, which yielded a unique sequence identifier tagged onto each individual sample library. Illumina-based sequencing yielded greater than 15,000 reads per sample. CLC Genomics Workbench version 12 (Qiagen) was used for OTU clustering and generation of abundance tables. Analyses were performed using the tutorial “OTU Clustering Step by Step” updated September 2, 2019 and available on the Qiagen website: https://resources.qiagenbioinformatics.com/tutorials/OTU_Clustering_Steps.pdf

### Microbial diversity and differential abundance analysis

Both alpha and beta diversity measures were calculated at the genus level using the vegan: Community Ecology Package in R (https://CRAN.R-project.org/package=vegan). Measures of alpha diversity included: the richness *S* (the number of taxa present in the community/sample), Chao 1 index 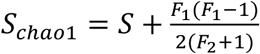, Shannon index 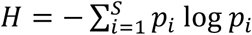, and evenness *J* = *H*/log*S*. Here, *F*_1_ and *F*_2_ are the count of singletons and doubletons, respectively, and *p_i_* is the relative abundance of taxon-*i* in the community. For beta diversity, we used the Bray-Curtis dissimilarity measure, which was also used in the Principal Coordinates Analysis (PCoA). We applied principal component analysis (PCA) on the expression levels of all immune markers based on the Euclidean distance.

Difference in microbiome compositions and immune expression levels by CDI status (i.e., different groups) and other covariates (i.e., age, sex, race and ethnicity) were tested by the permutational multivariate analysis of variance (PERMANOVA) using the “adonis” function in the vegan R package. All PERMANOVA tests were performed with the default 999 permutations based on the Bray-Curtis dissimilarity and Euclidean distance for microbial composition and immune marker data, respectively. Note that in the PERMANOVA tests, we only included subjects with known information of age, sex, race and ethnicity.

For differential abundance analysis, we used ANCOM^40^ (analysis of composition of microbiomes), with a Benjamini–Hochberg correction at 5% level of significance, and adjusted for age and sex. The Mann–Whitney U test was used to compare the difference of immune marker levels between different groups.

### Microbial correlation network analysis

The microbial correlation networks were constructed using SparCC ^41^ (sparse correlations for compositional data, https://github.com/luispedro/sparcc). Significant interactions were determined by the bootstrapped results (*N* = 100) using the script PseudoPvals in SparCC. Significant correlations with absolute sparse correlations ≥ 0.3 were visualized using Gephi (https://gephi.org/). We also used the NetShift^42^ (https://web.rniapps.net/netshift) to identify potential “driver” taxa underlying the differences of microbial correlation networks associated with CDI and Asymptomatic Carriage (or Non-CDI Diarrhea, and Non-CDI). The key driver taxa were identified based on the neighbor shift (NESH) score, Jaccard Index and delta betweenness (ΔB)^42^.

### Microbiome-Immune marker association analysis

Associations between the gut microbiota and host immune markers were quantified by Spearman correlation coefficients in combination with Benjamini-Hochberg FDR correction to account for multiple hypothesis testing (significance threshold *α ≤* 0.05). All included genera were required to be detected in ≥50% of all samples in each group.

### Classification with Random Forests model

To build a classification model capable of testing the overall contribution of immunological or microbial data in distinguishing the CDI status, we developed a multi-class random forests (RF) classifier. The data is split into a training set and a test set, with 70% of the data forming the training data and the remaining 30% forming the test set. The performance of the multi-class model was measured by micro-average and macro-average AUC. A macro-average score computed the metric independently for each group and then was averaged across all levels regardless of the number of samples in each group, whereas a micro-average will aggregate the contributions of all groups to compute the average metric.

To determine whether more specific host immune markers or gut microbial taxa could differentiate CDI subjects from Asymptomatic Carriage, Non-CDI Diarrhea and Non-CDI groups, we constructed the binary classifiers based on RF models with integrated immune markers and microbiome data. The performance of the classifiers were evaluated by a 5-fold cross validation. In order to reduce computation complexity and feature redundancy, a feature selection procedure was performed as follows. We first ranked all the features based on their mean decrease accuracy (MDA). Then we followed the “1-SE strategy” to select the minimum set of top features whose mean AUC is within one standard error of the mean AUC from the model with all of the features.

### Symbolic classification with genetic programming

Genetic programming (GP) is a genetic algorithm that searches the space of mathematical equations without any constraints on their forms^80^. GP involves reproduction, random mutation, crossover, a fitness function, and multiple generations of evolution of a population of computer programs to resolve a given task. GP is commonly used to investigate a functional relationship (i.e., a mathematical formula) between features in data (symbolic regression: SR) or to group data into categories (symbolic classification: SC). We employed Karoo GP^81^, a genetic programming application suite written in Python that support both SR and SC analysis, to derive simple formulas for CDI diagnosis. We performed a random data-split to create a training set (80% of the data) and a held-out test set (20% of the data) for ten times, which were used to evaluate the SC performance. Due to the different training sets, SC will derive different formulas, but their classification performances (in terms of Accuracy, Precision, Recall, F1-score) are quite comparable (Table S8). The formulas shown in Table 2 were derived based on the whole dataset. The Karoo GP was used with the following settings: (1) the fitness function (Kernel) is c (representing “classification”); (2) the type of tree is r (ramped half/half); (3) the maximum tree depth for the initial population is 6; (4) the number of trees per generation is 100; (5) the maximum number of generations is 190 (based on the converging results shown in Fig. S7); (6) constants include 0.1, 0.2, 0.3, 0.4 and 0.5; and (7) all other parameters are set as default values. The fitness function in SC is a maximization function, which will seek the highest fitness score among the trees in each generation. The sign of the final formula *f*(*i*) will be used for CDI diagnosis: the class of subject *i* is CDI if *f*(*i*) *>* 0; or Asymptomatic Carriage (or Non-CDI Diarrhea, Non-CDI) if *f*(*i*) *≤* 0.

To demonstrate the advantage of SC, for each classification task (i.e., CDI vs. Asymptomatic Carriage, CDI vs. Non-CDI Diarrhea, and CDI vs. Non-CDI), we also performed logistic regression (LR) using the same set of selected features as used in SC (Table 2). The LR models were constructed using the glm() function in R. The class of subject *i* is CDI if *p*(*i*) ≥ 0.5; or Asymptomatic Carriage (or Non-CDI Diarrhea, Non-CDI) if *p*(*i*) < 0.5.

## Data Availability

Data will be available from corresponding authors upon reasonable request.

## Acknowledgements

The authors thank all patients who participated in this study, as well as Carolyn Alonso, Javier Villafuerte Gálvez, and the technologists in the Beth Israel Deaconess Medical Center Clinical Microbiology Laboratory for their help with sample collection. The authors thank Zheng Sun for valuable discussion on the microbiome data analysis.

## Author contributions

Y.-Y.L, N.R.P., X.C., and C.P.K. conceived and designed the project. C.P.K., N.R.P., X.C. and K.D. performed the clinical study. X.C., H.X., and Q.L. contributed to the serum immune marker measurement. K.W.G. and A.J.G. performed fecal DNA extraction and bacterial 16S rRNA sequencing. S.K., X.-W.W., and Y.-Y.L. performed all the data analysis and wrote the manuscript. N.R.P., K.W.G., C.P.K., and K.D. edited the manuscript.

## Data and materials availability

Data will be available from corresponding authors upon reasonable request.

**Fig. S1.**
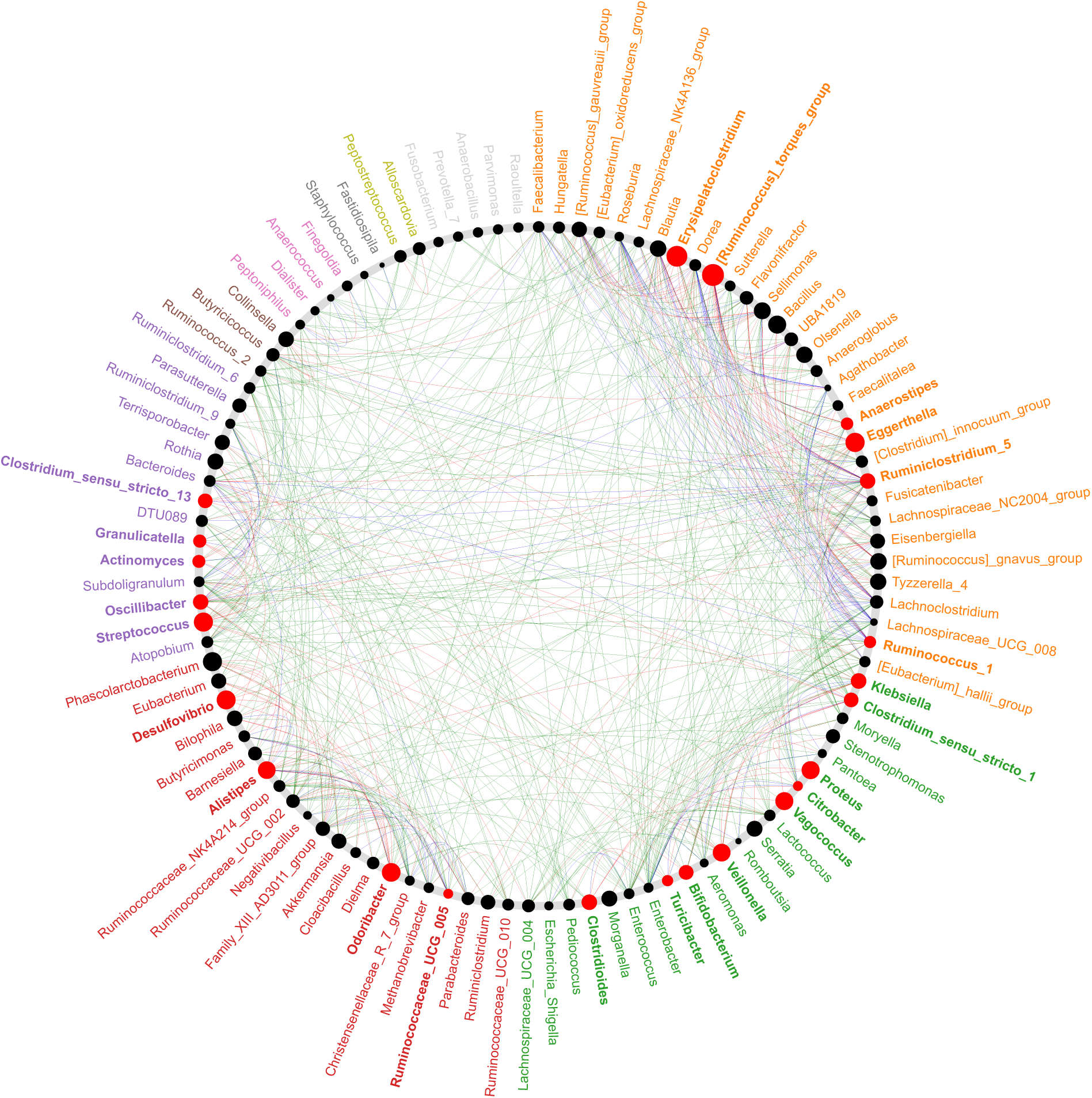
The “driver” taxa responsible for the change of microbial correlations between CDI and Asymptomatic Carriage. Node sizes are proportional to their scaled neighbor shift (NESH) score (i.e., a score identifying important microbial taxa of microbial association networks) and a node is colored red if its betweenness increases when comparing microbial correlation networks of CDI with that of Asymptomatic Carriage. All taxa belonging to same community (common sub-network) are randomly assigned a color to their labels. Red (or green) edges represent microbial correlations that are only present in the CDI (or Asymptomatic Carriage) network, respectively. Blue edges present common microbial correlations that are present in both networks.

**Fig. S2.**
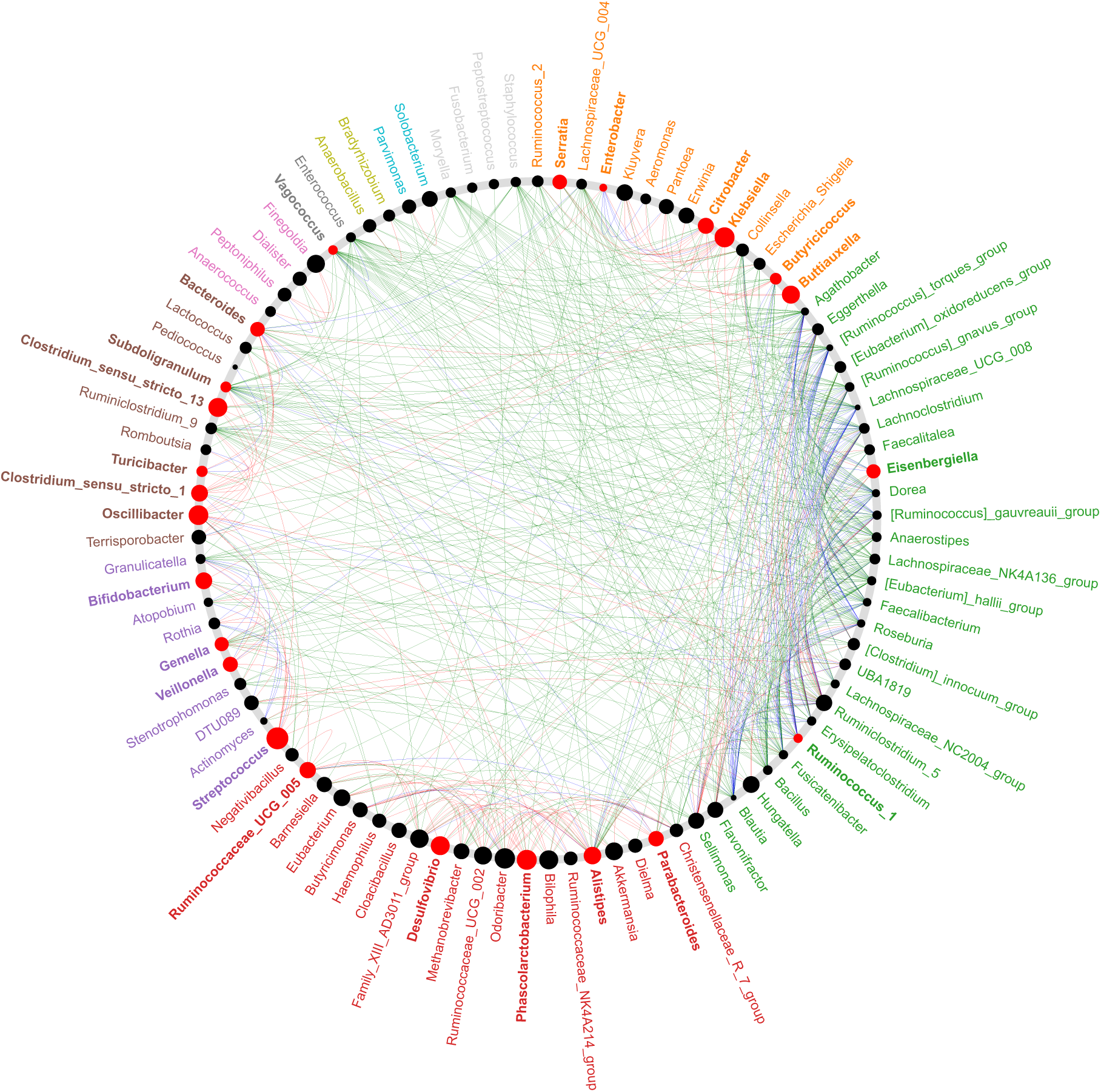
The “driver” taxa responsible for the change of microbial correlations between CDI and Non-CDI Diarrhea. Node sizes are proportional to their scaled NESH score and a node is colored red if its betweenness increases when comparing microbial correlation networks of CDI with that of Non-CDI Diarrhea. All taxa belonging to same community (common subnetwork) are randomly assigned a color to their labels. Red (or green) edges represent microbial correlations that are only present in the CDI (or Non-CDI Diarrhea) network, respectively. Blue edges present common microbial correlations that are present in both networks.

**Fig. S3.**
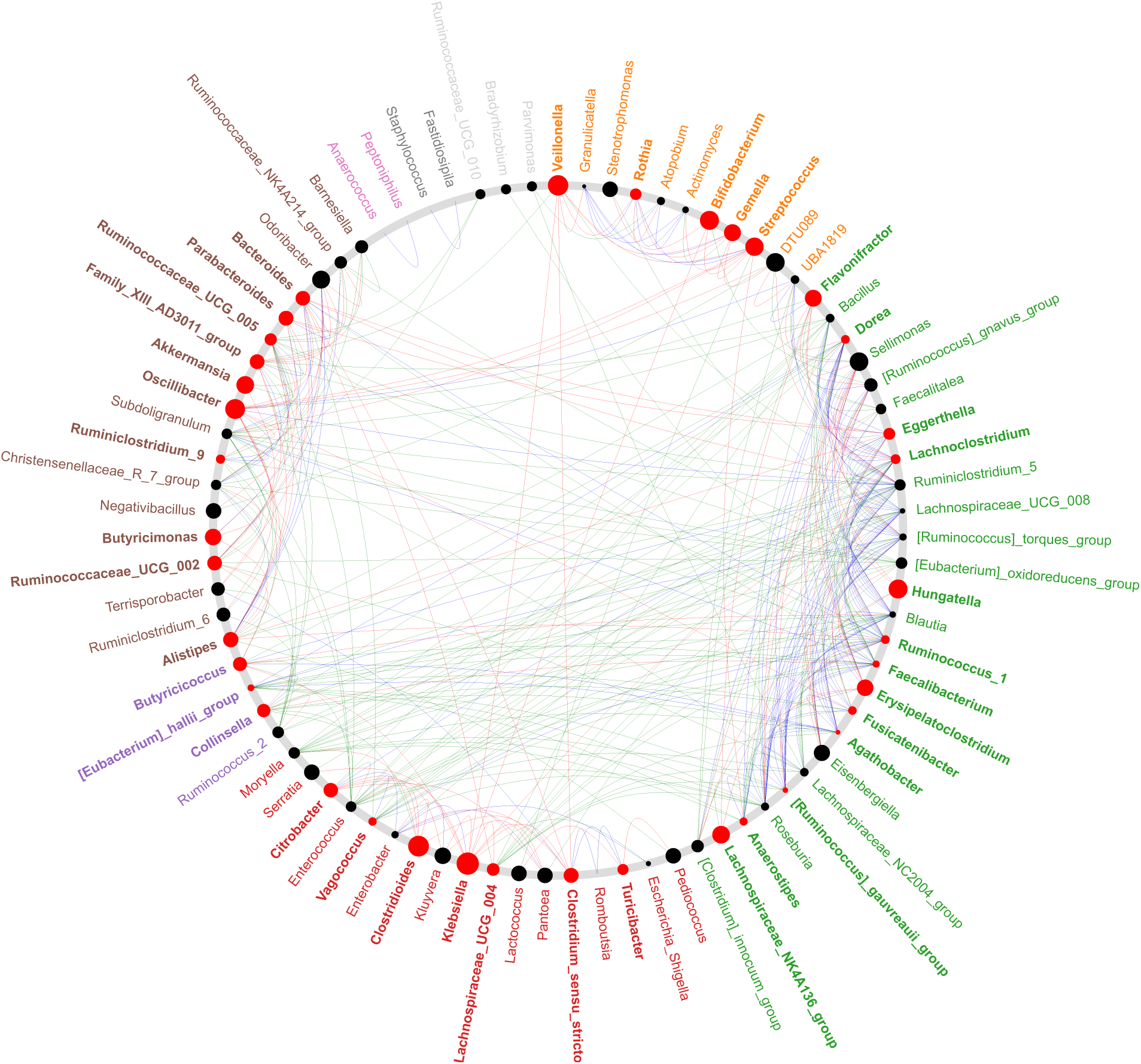
The potential “driver taxa” responsible for the change of microbial correlations between CDI and Non-CDI. Node sizes are proportional to their scaled NESH score and a node is colored red if its betweenness increases when comparing microbial correlation networks of CDI with that of Non-CDI. All taxa belonging to same community (common sub-network) are randomly assigned a color to their labels. Red (or green) edges represent microbial correlations that are only present in the CDI (or Non-CDI) network, respectively. Blue edges present common microbial correlations that are present in both networks.

**Fig. S4.**
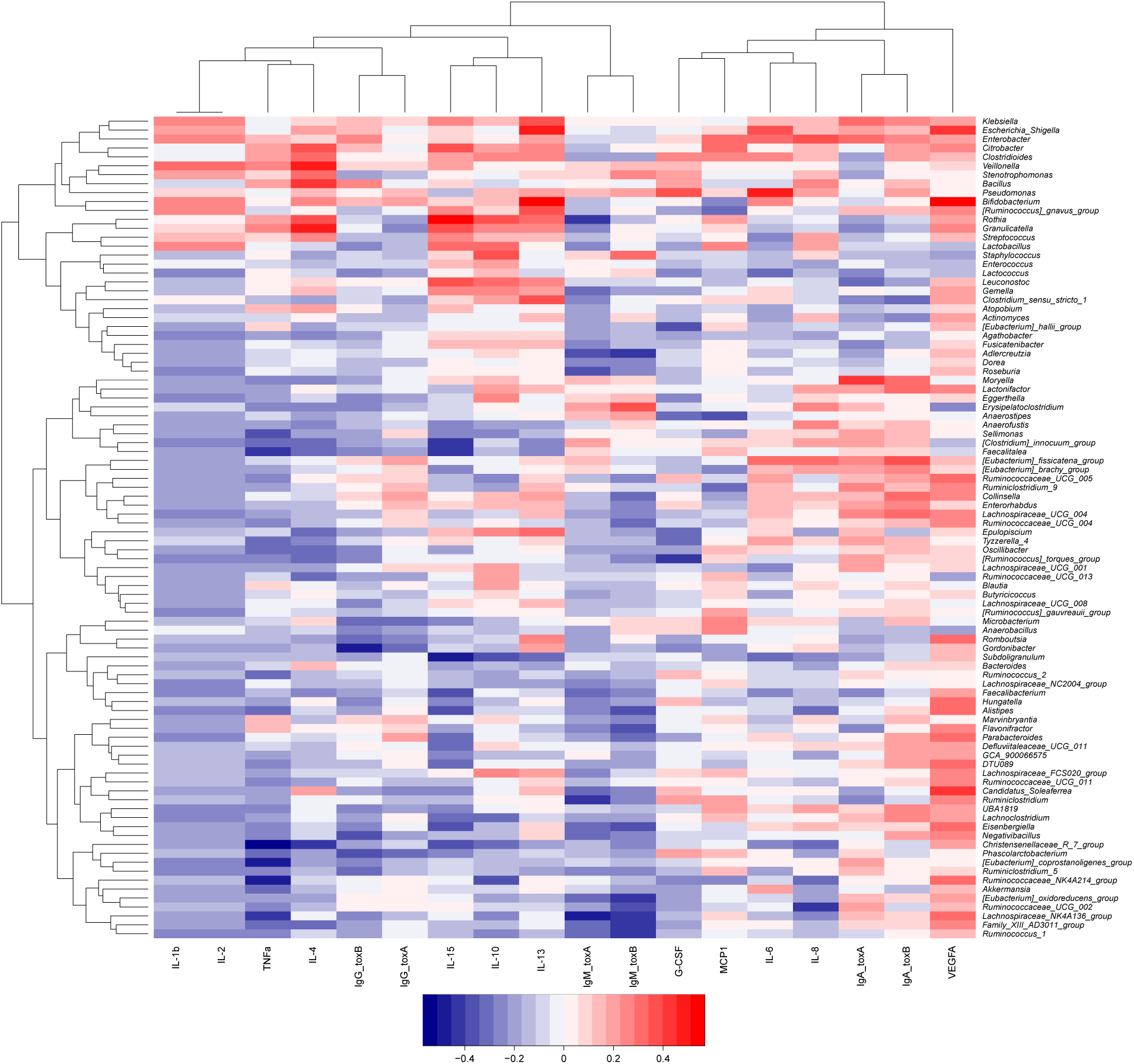
Significant correlations between gut microbial abundances and host immune markers in the Control group. Gut microbial compositions and host immune markers were clustered through hierarchical clustering. Rows correspond to bacterial taxa at genus level; columns correspond to host immune markers. Red/blue represents positive/negative association, respectively. The intensity of the colors denotes the strength of correlation between the genus abundance and the immunological expression level.

**Fig. S5.**
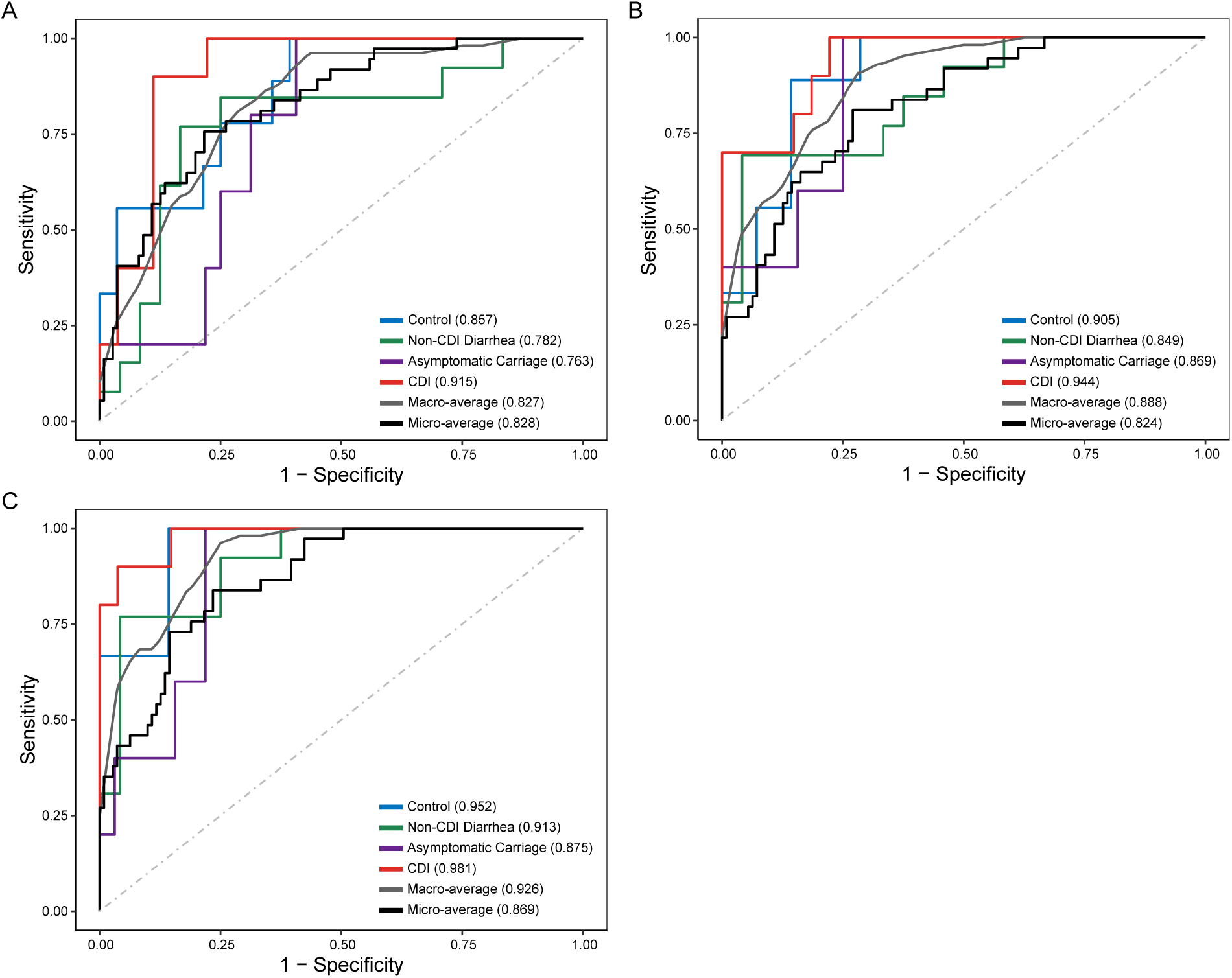
Gut microbiota and host immune markers can accurately differentiate different groups in multi-class classification models. (**A**) Use host immune markers alone. (**B**) Use gut microbiota data (at genus level) alone. (**C**) The integration of host immune markers and microbial data. The performance of each classifier is measured by the macro-average and microaverage AUCs.

**Fig. S6.**
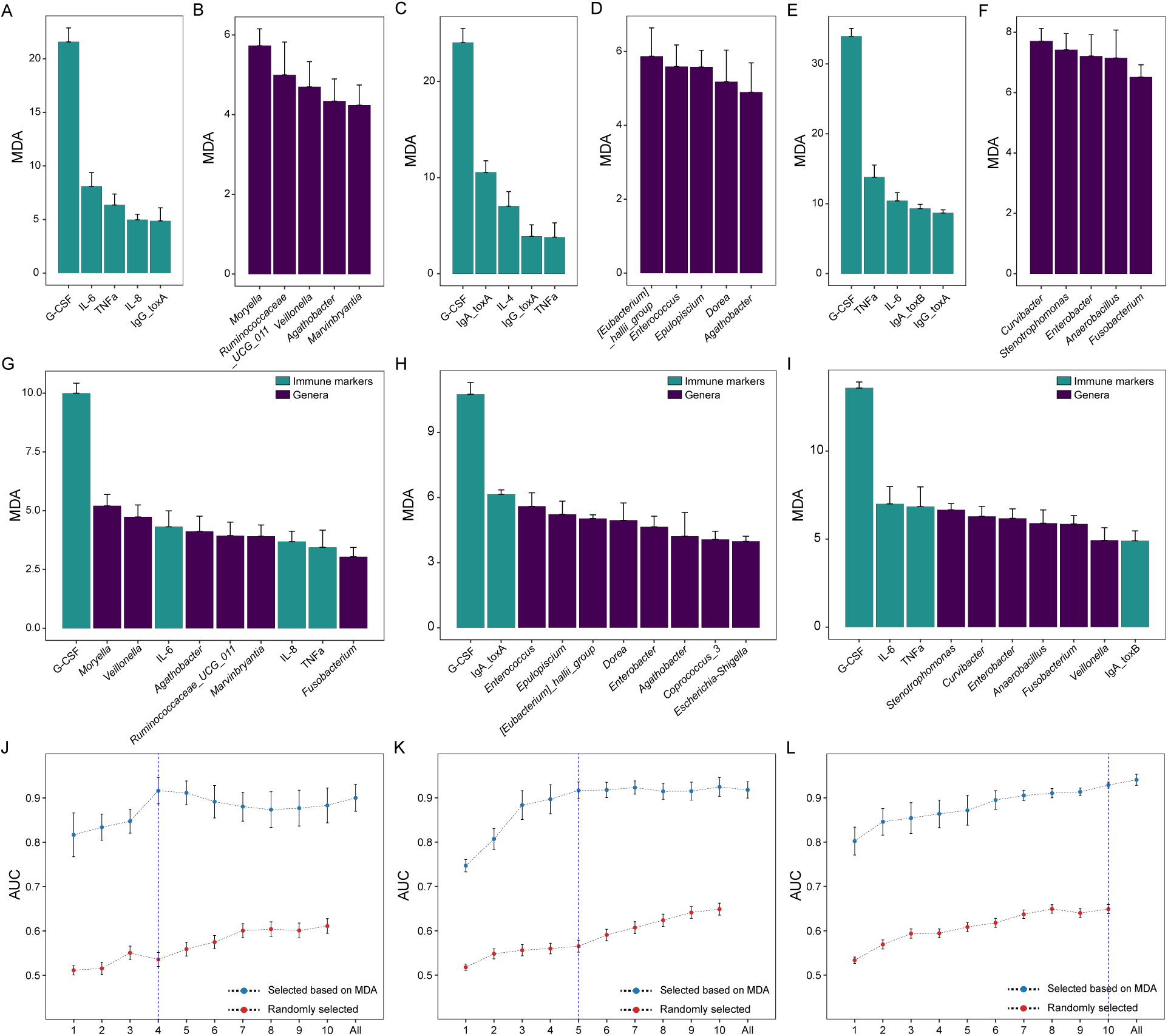
Using the mean decrease accuracy (MDA) ranking and the 1-SE rule to select features to distinguish CDI from other groups. The most important features of cytokine data, microbiome data, and the integration of cytokines and microbiome data in classifying CDI vs. Asymptomatic Carriage (**A, B and G**), CDI vs. Non-CDI Diarrhea (**C, D and H**) and CDI vs. Non-CDI (**E, F and I**). The performance of classifiers using different sets of integrated features: selected based on MDA or randomly selected in CDI vs Asymptomatic Carriage (**J**), CDI vs Non-CDI Diarrhea (**K**) and CDI vs Non-CDI (**L**). The minimum set of features selected based on the MDA ranking and the 1-SE rule is highlighted by a vertical blue dashed line. Error bars represent the standard errors of the means (SEM).

**Fig. S7.**
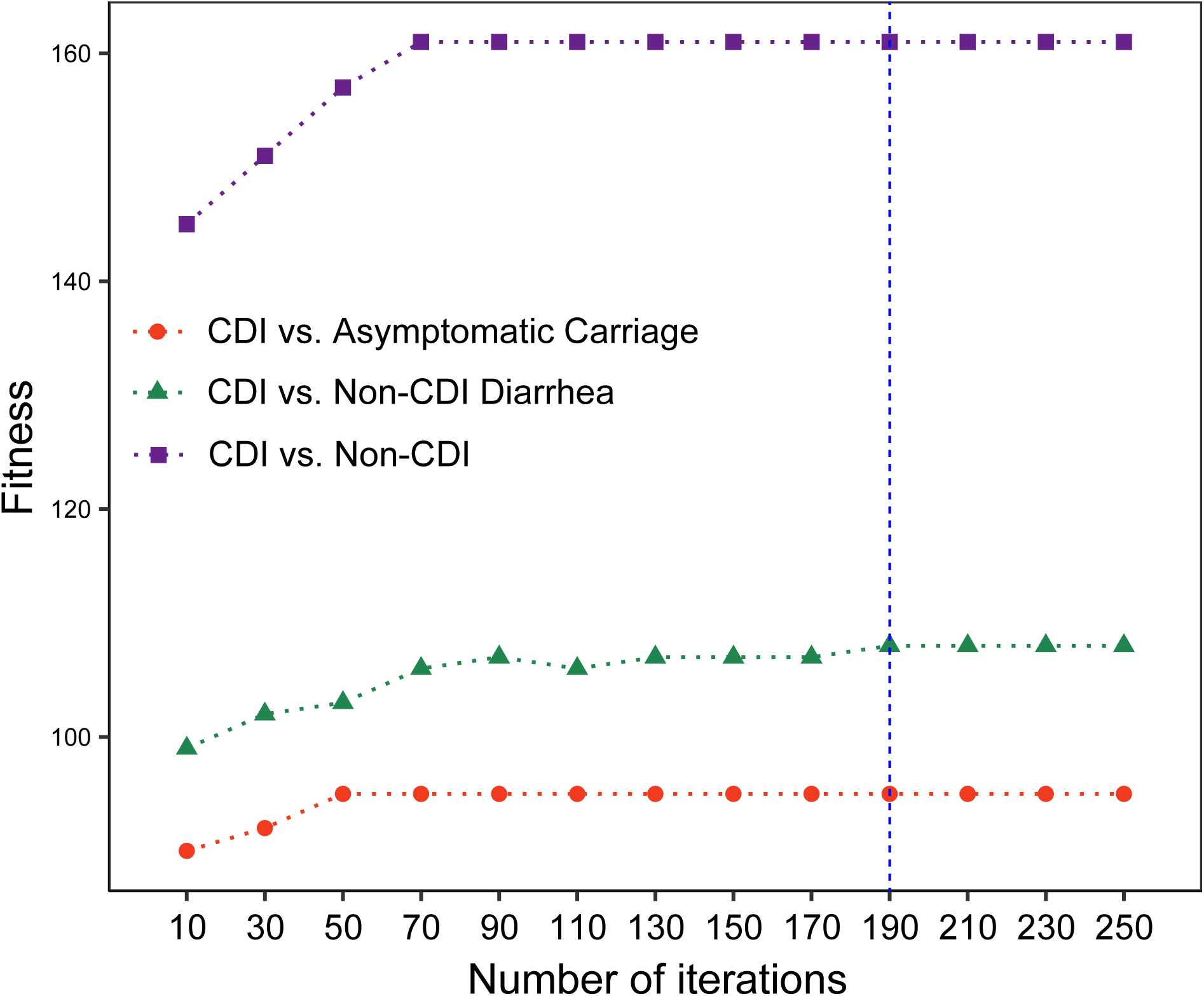
The fitness evolution during the symbolic classification based on genetic programming. The fitness function is a maximization function, and the tree with highest fitness score in each iteration were plotted. The final selected number of generations is highlighted with a vertical blue dashed line.

**Table S1.** Sample sizes of different data types in different groups.

| **Characteristics** | **NAAT negative** |  | **NAAT positive** |  | **Total** |
| --- | --- | --- | --- | --- | --- |
|  | **Control** | **Non-CDI**<br>**Diarrhea** | **Asymptomatic**<br>**Carriage** | **CDI** |  |
| Immunological data | 45 | 44 | 35 | 99 | 223 |
| Microbial data | 41 | 42 | 33 | 91 | 207 |
| Immunological &<br>microbial data | 39 | 42 | 28 | 78 | 187 |

**Table S2.** Permutational multivariate analysis of variance (PERMANOVA) in microbial compositions and immune markers. CDI statuses: Control, Non-CDI Diarrhea, Asymptomatic Carriage, and CDI. Race: White, Native American, Asian, African American, Pacific Islander and mixed origin. Ethnicity: Hispanic and Not Hispanic. Here F represents the F-statistic: a larger F value indicate that the between-group variation is greater than within-group variation. R^2^ represents the variation explained by the model. P represents the *P*-value calculated from permutation.

|  | **Microbiome** |  |  | **Cytokines** |  |  |
| --- | --- | --- | --- | --- | --- | --- |
| Test factors | F | R<sup>2</sup> | P | F | R<sup>2</sup> | P |
| CDI status | 2.285 | 0.0388 | 0.001 | 3.351 | 0.052 | 0.016 |
| Age | 1.605 | 0.009 | 0.081 | 0.541 | 0.003 | 0.516 |
| Sex | 1.557 | 0.009 | 0.095 | 0.916 | 0.005 | 0.372 |
| Race | 0.881 | 0.031 | 0.832 | 1.595 | 0.050 | 0.153 |
| Ethnicity | 0.476 | 0.003 | 0.961 | 0.206 | 0.001 | 0.771 |

**Table S3.**
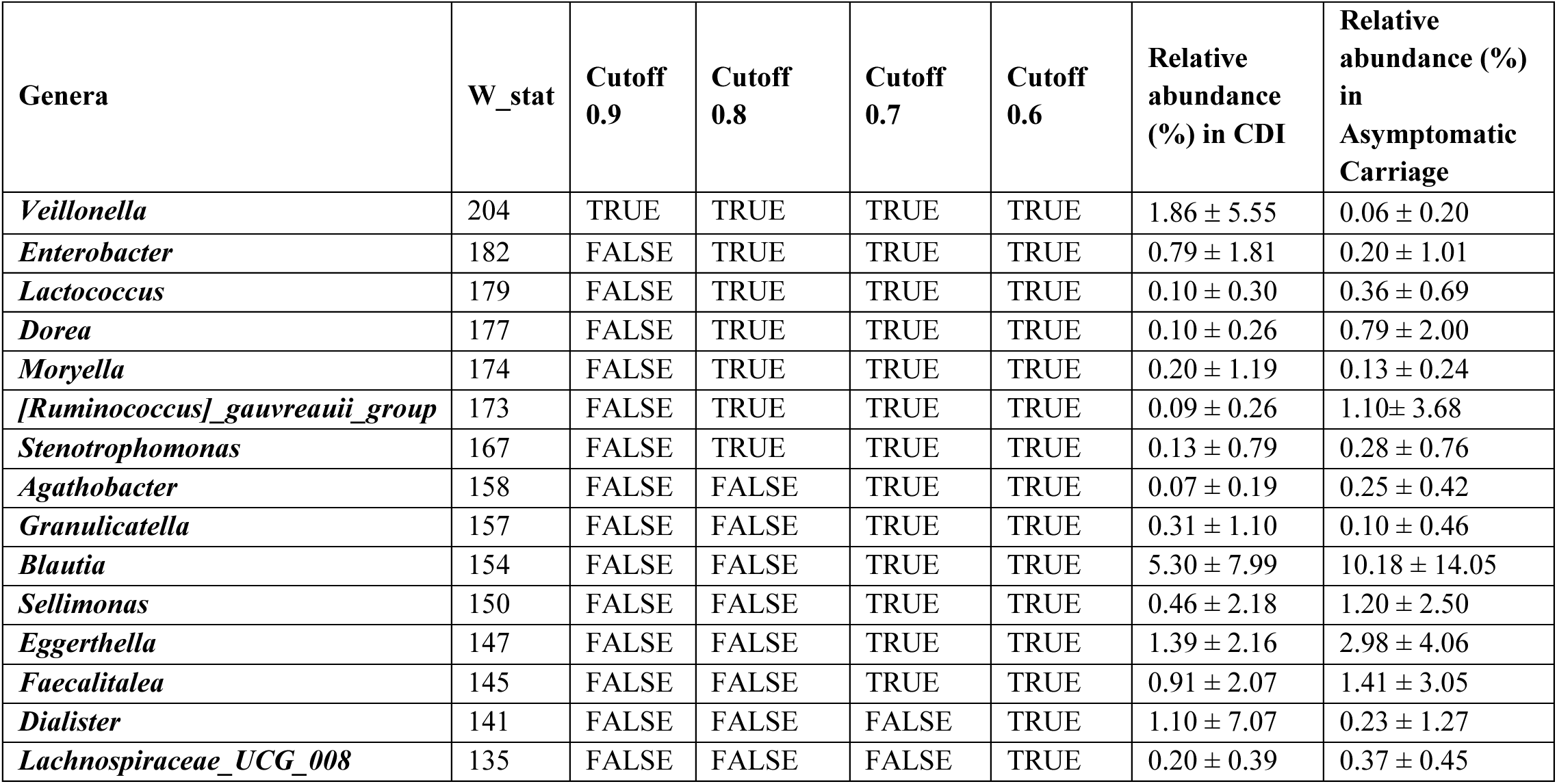
Differentially abundant genera between CDI and Asymptomatic Carriage groups detected by ANCOM, adjusted for age and sex. For each genus, the first column represents its W statistic, and subsequent four columns represent logical indicators of whether it is differentially abundant under a series of cutoffs (0.9, 0.8, 0.7 and 0.6). The last two columns represent its relative abundance (mean ± standard deviation) in the two groups.

**Table S4.** Differentially abundant genera between CDI and Non-CDI Diarrhea groups detected by ANCOM, adjusted for age and sex. For each genus, the first column represents its W statistic, and subsequent four columns represent logical indicators of whether it is differentially abundant under a series of cutoffs (0.9, 0.8, 0.7 and 0.6). The last two columns represent its relative abundance (mean ± standard deviation) in the two groups.

| Genera | W_stat | detected_0.9 | detected_0.8 | detected_0.7 | detected_0.6 | Relative abundance (%) in CDI | Relative abundance (%) in Non-CDI Diarrhea |
| --- | --- | --- | --- | --- | --- | --- | --- |
| *Clostridioides* | 206 | TRUE | TRUE | TRUE | TRUE | 0.67 ± 1.80 | 0.03 ± 0.16 |
| *[Eubacterium]_hallii_group* | 199 | TRUE | TRUE | TRUE | TRUE | 0.40 ± 2.12 | 1.14 ± 2.05 |
| *Collinsella* | 195 | TRUE | TRUE | TRUE | TRUE | 0.57 ± 1.59 | 2.57 ± 5.33 |
| *Enterobacter* | 189 | TRUE | TRUE | TRUE | TRUE | 0.79 ± 1.81 | 0.05 ± 0.20 |
| *Epulopiscium* | 166 | FALSE | TRUE | TRUE | TRUE | 0.06 ± 0.30 | 0.00 ± 0.01 |
| *Agathobacter* | 165 | FALSE | TRUE | TRUE | TRUE | 0.07 ± 0.19 | 0.42 ± 0.93 |
| *Dorea* | 165 | FALSE | TRUE | TRUE | TRUE | 0.10 ± 0.26 | 0.96 ± 2.24 |
| *Escherichia_Shigella* | 163 | FALSE | FALSE | TRUE | TRUE | 3.54 ± 6.46 | 1.84 ± 5.01 |
| *Eisenbergiella* | 149 | FALSE | FALSE | TRUE | TRUE | 1.03 ± 3.36 | 0.10 ± 0.40 |
| *Stenotrophomonas* | 147 | FALSE | FALSE | TRUE | TRUE | 0.13 ± 0.79 | 0.09 ± 0.17 |
| *Streptococcus* | 147 | FALSE | FALSE | TRUE | TRUE | 6.16 ± 13.27 | 7.00 ± 7.39 |
| *Dialister* | 138 | FALSE | FALSE | FALSE | TRUE | 1.10 ± 7.07 | 0.06 ± 0.25 |
| *Ruminiclostridium* | 137 | FALSE | FALSE | FALSE | TRUE | 0.08 ± 0.44 | 0.00 ± 0.01 |
| *Fusobacterium* | 131 | FALSE | FALSE | FALSE | TRUE | 0.18 ± 0.51 | 0.01 ± 0.04 |
| *Klebsiella* | 131 | FALSE | FALSE | FALSE | TRUE | 1.75 ± 6.94 | 0.58 ± 2.74 |
| *Veillonella* | 125 | FALSE | FALSE | FALSE | TRUE | 1.86 ± 5.55 | 0.27 ± 0.78 |

**Table S5.** Differentially abundant genera between CDI and Non-CDI groups detected by ANCOM, adjusted for age and sex. For each genus, the first column represents its W statistic, and subsequent four columns represent logical indicators of whether it is differentially abundant under a series of cutoffs (0.9, 0.8, 0.7 and 0.6). The last two columns represent its relative abundance (mean ± standard deviation) in the two groups.

| Genera | W_stat | detected<br>_0.9 | detected_<br>0.8 | detected_<br>0.7 | detected_<br>0.6 | Relative<br>abundance<br>(%) in CDI | Relative<br>abundance (%)<br>in Non-CDI |
| --- | --- | --- | --- | --- | --- | --- | --- |
| *Clostridioides* | 201 | TRUE | TRUE | TRUE | TRUE | 0.67 ± 1.81 | 0.08 ± 0.26 |
| *Veillonella* | 201 | TRUE | TRUE | TRUE | TRUE | 1.86 ± 5.58 | 0.14 ± 0.50 |
| *Enterobacter* | 200 | TRUE | TRUE | TRUE | TRUE | 0.79 ± 1.82 | 0.08 ± 0.55 |
| *Klebsiella* | 196 | TRUE | TRUE | TRUE | TRUE | 1.75 ± 6.98 | 0.49 ± 2.25 |
| *Collinsella* | 194 | TRUE | TRUE | TRUE | TRUE | 0.57 ± 1.60 | 2.29 ± 4.64 |
| *Fusobacterium* | 194 | TRUE | TRUE | TRUE | TRUE | 0.18 ± 0.51 | 0.04 ± 0.25 |
| *[Eubacterium]_hallii_group* | 193 | TRUE | TRUE | TRUE | TRUE | 0.40 ± 2.13 | 1.21 ± 2.55 |
| *Stenotrophomonas* | 193 | TRUE | TRUE | TRUE | TRUE | 0.13 ± 0.79 | 0.30 ± 1.06 |
| *Escherichia_Shigella* | 191 | TRUE | TRUE | TRUE | TRUE | 3.54 ± 6.50 | 1.96 ± 5.31 |
| *[Ruminococcus]_gnavus_group* | 185 | FALSE | TRUE | TRUE | TRUE | 1.73 ± 3.15 | 0.50 ± 1.81 |
| *Agathobacter* | 179 | FALSE | TRUE | TRUE | TRUE | 0.07 ± 0.19 | 0.50 ± 1.37 |
| *Dialister* | 176 | FALSE | TRUE | TRUE | TRUE | 1.10 ± 7.11 | 0.10 ± 0.69 |
| *Dorea* | 170 | FALSE | TRUE | TRUE | TRUE | 0.10 ± 0.26 | 0.86 ± 2.17 |
| *Lactococcus* | 169 | FALSE | TRUE | TRUE | TRUE | 0.10 ± 0.30 | 0.23 ± 0.56 |
| *Anaerobacillus* | 164 | FALSE | FALSE | TRUE | TRUE | 0.02 ± 0.06 | 0.00 ± 0.01 |
| *Moryella* | 164 | FALSE | FALSE | TRUE | TRUE | 0.20 ± 1.19 | 0.11 ± 0.26 |
| *Adlercreutzia* | 162 | FALSE | FALSE | TRUE | TRUE | 0.00 ± 0.02 | 0.08 ± 0.33 |
| *Family_XIII_AD3011_group* | 161 | FALSE | FALSE | TRUE | TRUE | 0.03 ± 0.07 | 0.09 ± 0.16 |
| *Erysipelatoclostridium* | 160 | FALSE | FALSE | TRUE | TRUE | 3.56 ± 7.56 | 0.80 ± 1.79 |
| *[Eubacterium]_brachy_group* | 154 | FALSE | FALSE | TRUE | TRUE | 0.01 ± 0.03 | 0.04 ± 0.10 |
| *Campylobacter* | 154 | FALSE | FALSE | TRUE | TRUE | 0.03 ± 0.11 | 0.00 ± 0.00 |
| *Citrobacter* | 154 | FALSE | FALSE | TRUE | TRUE | 0.41 ± 2.11 | 0.20 ± 1.09 |
| *Clostridium_sensu_stricto_1* | 151 | FALSE | FALSE | TRUE | TRUE | 0.72 ± 1.76 | 0.38 ± 1.19 |
| *Clostridium_sensu_stricto_13* | 151 | FALSE | FALSE | TRUE | TRUE | 0.07 ± 0.41 | 0.00 ± 0.01 |
| *Akkermansia* | 149 | FALSE | FALSE | TRUE | TRUE | 3.3 ± 9.68 | 6.41 ± 12.03 |
| *Alistipes* | 142 | FALSE | FALSE | FALSE | TRUE | 1.28 ± 3.05 | 1.30 ± 2.34 |
| *Bacillus* | 139 | FALSE | FALSE | FALSE | TRUE | 0.01 ± 0.03 | 0.00 ± 0.01 |
| *Enterorhabdus* | 137 | FALSE | FALSE | FALSE | TRUE | 0.01 ± 0.03 | 0.08 ± 0.55 |
| *Pantoea* | 137 | FALSE | FALSE | FALSE | TRUE | 0.01 ± 0.03 | 0.00 ± 0.00 |
| *Ruminococcaceae_UCG_004* | 134 | FALSE | FALSE | FALSE | TRUE | 0.05 ± 0.17 | 0.15 ± 0.41 |
| *Curvibacter* | 132 | FALSE | FALSE | FALSE | TRUE | 0.01 ± 0.01 | 0.00 ± 0.00 |
| *Granulicatella* | 131 | FALSE | FALSE | FALSE | TRUE | 0.31 ± 1.11 | 0.08 ± 0.27 |
| *Lachnospiraceae_NC2004_group* | 131 | FALSE | FALSE | FALSE | TRUE | 0.02 ± 0.04 | 0.05 ± 0.09 |
| *Ruminiclostridium_5* | 131 | FALSE | FALSE | FALSE | TRUE | 0.50 ± 1.11 | 1.33 ± 2.79 |
| *Epulopiscium* | 130 | FALSE | FALSE | FALSE | TRUE | 0.06 ± 0.30 | 0.01 ± 0.04 |
| *Robinsoniella* | 130 | FALSE | FALSE | FALSE | TRUE | 0.06 ± 0.33 | 0.01 ± 0.07 |
| *[Eubacterium]_coprostanoligenes_group* | 128 | FALSE | FALSE | FALSE | TRUE | 0.13 ± 0.39 | 0.39 ± 2.19 |
| *Eggerthella* | 128 | FALSE | FALSE | FALSE | TRUE | 1.39 ± 2.17 | 1.86 ± 2.96 |
| *Erwinia* | 126 | FALSE | FALSE | FALSE | TRUE | 0.00 ± 0.00 | 0.00 ± 0.00 |
| *[Ruminococcus]_gauvreauii_group* | 124 | FALSE | FALSE | FALSE | TRUE | 0.09 ± 0.26 | 0.45 ± 2.04 |

**Table S6.** Characteristics of microbial correlation networks associated with different groups.

| **Groups** | **Average degree** | **Clustering coefficient** | **Edges** | **Graph density** | **Modularity** | **Nodes** |
| --- | --- | --- | --- | --- | --- | --- |
| **Control** | 9.475 | 0.368 | 938 | 0.048 | 0.377 | 198 |
| **Non-CDI Diarrhea** | 11.314 | 0.474 | 1171 | 0.055 | 0.271 | 207 |
| **Asymptomatic Carriage** | 9.730 | 0.349 | 973 | 0.049 | 0.442 | 200 |
| **CDI** | 5.200 | 0.502 | 299 | 0.046 | 0.568 | 115 |

**Table S7.** Comparison of host immune markers in different groups. Mean (Q1, Q3); p-value calculated with Mann-Whitney U test.

| Immune markers | Control (n=45) | Non-CDI Diarrhea (n=44) | Asymptomatic Carriage (n=35) | CDI (n=99) | P-value (CDI vs. Asymptomatic Carriage) | P-value (CDI vs. Non-CDI Diarrhea) | P-value (CDI vs. Non-CDI) |
| --- | --- | --- | --- | --- | --- | --- | --- |
| IgA_toxA | 33.63 (7.24, 62.54) | 16.94 (8.44, 16.96) | 44.59 (10.21, 102.50) | 48.41 (12.10, 104) | 0.543 | < 0.001 | 0.001 |
| IgG_toxA | 19.87 (9.75, 22.43) | 24.20 (11.11, 27.77) | 22.20 (11.48, 24.86) | 40.09 (14.77, 59.18) | 0.009 | 0.002 | < 0.001 |
| IgM_toxA | 2.04 (0.00, 2.87) | 2.45 (0.00, 3.36) | 2.15 (0.00, 2.98) | 2.09 (0.00, 2.84) | 0.316 | 0.643 | 0.172 |
| IgA_toxB | 9.73 (2.68, 8.61) | 18.78 (4.07, 19.18) | 23.02 (5.80, 20.12) | 35.07 (4.76, 67.79) | 0.469 | 0.102 | 0.002 |
| IgG_toxB | 9.66 (4.30, 9.80) | 11.97 (5.92, 13.46) | 13.57 (3.98, 15.98) | 14.61 (4.92, 17.56) | 0.980 | 0.870 | 0.379 |
| IgM_toxB | 12.12 (2.28, 9.24) | 13.72 (2.78, 12.77) | 11.08 (2.74, 9.04) | 14.99 (1.87, 10.47) | 0.261 | 0.192 | 0.256 |
| GCSF | 11.27 (0.46, 14.17) | 49.37 (2.18, 29.36) | 20.01 (2.18, 20.49) | 386.64 (22.56, 159.95) | < 0.001 | < 0.001 | < 0.001 |
| IL-10 | 8.29 (0.00, 3.15) | 33.22 (0.00, 14.06) | 9.05 (0.00, 9.78) | 35.17 (1.63, 27.99) | 0.002 | 0.021 | < 0.001 |
| IL-13 | 1.38 (0.00, 0.00) | 1.97 (0.00, 0.34) | 5.22 (0.00, 0.00) | 9.35 (0.00, 1.09) | 0.529 | 0.593 | 0.167 |
| IL-15 | 1.56 (0.00, 0.24) | 2.82 (0.00, 3.28) | 2.03 (0.00, 1.46) | 5.22 (0.19, 5.33) | 0.007 | 0.037 | < 0.001 |
| IL-1b | 0.05 (0.00, 0.00) | 0.11 (0.00, 0.00) | 0.44 (0.00, 0.00) | 0.7 (0.00, 0.00) | 0.920 | 0.387 | 0.159 |
| IL-2 | 0.04 (0.00, 0.00) | 0.05 (0.00, 0.00) | 0.45 (0.00, 0.00) | 1.4 (0.00, 0.00) | 0.630 | 0.151 | 0.051 |
| IL-4 | 1.88 (0.00, 0.00) | 9.58 (2.57, 12.44) | 5.73 (0.00, 0.00) | 11.54 (0.00, 9.12) | 0.002 | 0.011 | 0.032 |
| IL-6 | 15.78 (0.00, 3.77) | 24.97 (0.00, 10.71) | 9.52 (0.00, 5.46) | 47.09 (2.52, 37.71) | < 0.001 | < 0.001 | < 0.001 |
| IL-8 | 100.51 (12.37, 59.19) | 80.85 (15.22, 73.45) | 59.78 (10.33, 44.76) | 128.05 (27.20, 122.24) | < 0.001 | 0.004 | < 0.001 |
| MCP1 | 477.00 (397.19, 545.00) | 591.61 (399.55, 779.32) | 613.28 (430.85, 791.37) | 844.96 (522.41, 990.98) | 0.053 | 0.020 | < 0.001 |
| TNFa | 8.61 (6.20, 10.95) | 21.76 (8.93, 21.22) | 12.45 (4.91, 14.84) | 26.68 (13.88, 28.94) | < 0.001 | 0.006 | < 0.001 |
| VEGFA | 102.78 (35.76, 118.27) | 109.85 (34.53, 127.69) | 118.44 (27.54, 188.60) | 125.93 (19.54, 140.49) | 0.499 | 0.603 | 0.390 |

**Table S8.** Accuracy, Precision, Recall and F1-score of symbolic classification in CDI diagnosis. CDI subjects were considered as either true positive or true negative. Results shown in the parenthesis represents the latter case. The performance of the symbolic classification model evaluated by cross-validation. We randomly split the dataset to form a training set (80% of the data) and a test set (20% of the data) in 10 different ways. Each time, for each classification task (diagnostic goal), we learned the SC model from the training dataset and evaluated it on the test dataset. Data represents as mean ± standard deviation.

| Diagnostic goal | Accuracy | Precision | Recall | F1-score |
| --- | --- | --- | --- | --- |
| CDI vs. Asymptomatic Carriage | $0.873 \pm 0.085$ | $0.878 \pm 0.101$<br>($0.857 \pm 0.180$) | $0.963 \pm 0.043$<br>($0.639 \pm 0.229$) | $0.915 \pm 0.061$<br>($0.718 \pm 0.185$) |
| CDI vs. Non-CDI Diarrhea | $0.883 \pm 0.036$ | $0.899 \pm 0.078$<br>($0.865 \pm 0.098$) | $0.912 \pm 0.070$<br>($0.847 \pm 0.110$) | $0.901 \pm 0.039$<br>($0.846 \pm 0.055$) |
| CDI vs. Non-CDI | $0.818 \pm 0.040$ | $0.770 \pm 0.102$<br>($0.861 \pm 0.059$) | $0.784 \pm 0.120$<br>($0.839 \pm 0.091$) | $0.769 \pm 0.074$<br>($0.845 \pm 0.036$) |

## REFERENCES AND NOTES

1 Lessa, F. C. et al. Burden of Clostridium difficile infection in the United States. N Engl J Med 372, 825–834, doi:10.1056/NEJMoa1408913 (2015).

2 Depestel, D. D. & Aronoff, D. M. Epidemiology of Clostridium difficile infection. J Pharm Pract 26, 464–475, doi:10.1177/0897190013499521 (2013).

3 McDonald, L. C. et al. Clinical Practice Guidelines for Clostridium difficile Infection in Adults and Children: 2017 Update by the Infectious Diseases Society of America (IDSA) and Society for Healthcare Epidemiology of America (SHEA). Clin Infect Dis 66, e1-e48, doi:10.1093/cid/cix1085 (2018).

4 Bagdasarian, N., Rao, K. & Malani, P. N. Diagnosis and treatment of Clostridium difficile in adults: a systematic review. JAMA 313, 398–408, doi:10.1001/jama.2014.17103 (2015).

5 Rineh, A., Kelso, M. J., Vatansever, F., Tegos, G. P. & Hamblin, M. R. Clostridium difficile infection: molecular pathogenesis and novel therapeutics. Expert Rev Anti Infect Ther 12, 131–150, doi:10.1586/14787210.2014.866515 (2014).

6 Stevens, V., Dumyati, G., Fine, L. S., Fisher, S. G. & van Wijngaarden, E. Cumulative antibiotic exposures over time and the risk of Clostridium difficile infection. Clin Infect Dis 53, 42–48, doi:10.1093/cid/cir301 (2011).

7 Slimings, C. & Riley, T. V. Antibiotics and hospital-acquired Clostridium difficile infection: update of systematic review and meta-analysis. J Antimicrob Chemother 69, 881–891, doi:10.1093/jac/dkt477 (2014).

8 Lewis, B. B. et al. Loss of Microbiota-Mediated Colonization Resistance to Clostridium difficile Infection With Oral Vancomycin Compared With Metronidazole. J Infect Dis 212, 1656–1665, doi:10.1093/infdis/jiv256 (2015).

9 Becattini, S., Taur, Y. & Pamer, E. G. Antibiotic-Induced Changes in the Intestinal Microbiota and Disease. Trends Mol Med 22, 458–478, doi:10.1016/j.molmed.2016.04.003 (2016).

10 Buffie, C. G. et al. Profound alterations of intestinal microbiota following a single dose of clindamycin results in sustained susceptibility to Clostridium difficile-induced colitis. Infect Immun 80, 62–73, doi:10.1128/IAI.05496-11 (2012).

11 Perez-Cobas, A. E. et al. Structural and functional changes in the gut microbiota associated to Clostridium difficile infection. Front Microbiol 5, 335, doi:10.3389/fmicb.2014.00335 (2014).

12 Theriot, C. M. et al. Antibiotic-induced shifts in the mouse gut microbiome and metabolome increase susceptibility to Clostridium difficile infection. Nat Commun 5, 3114, doi:10.1038/ncomms4114 (2014).

13 Leffler, D. A., & Lamont, J. T. Clostridium difficile Infection. N Engl J Med 373, 287–288, doi:10.1056/NEJMc1506004 (2015).

14 Genth, H., Dreger, S. C., Huelsenbeck, J. & Just, I. Clostridium difficile toxins: more than mere inhibitors of Rho proteins. Int J Biochem Cell Biol 40, 592–597, doi:10.1016/j.biocel.2007.12.014 (2008).

15 Sun, X., He, X., Tzipori, S., Gerhard, R. & Feng, H. Essential role of the glucosyltransferase activity in Clostridium difficile toxin-induced secretion of TNF-alpha by macrophages. Microb Pathog 46, 298–305, doi:10.1016/j.micpath.2009.03.002 (2009).

16 Riegler, M. et al. Clostridium difficile toxin B is more potent than toxin A in damaging human colonic epithelium in vitro. J Clin Invest 95, 2004–2011, doi:10.1172/JCI117885 (1995).

17 Sun, X. & Hirota, S. A. The roles of host and pathogen factors and the innate immune response in the pathogenesis of Clostridium difficile infection. Mol Immunol 63, 193–202, doi:10.1016/j.molimm.2014.09.005 (2015).

18 Kelly, C. P. & Kyne, L. The host immune response to Clostridium difficile. J Med Microbiol 60, 1070–1079, doi:10.1099/jmm.0.030015-0 (2011).

19 Bibbo, S. et al. Role of microbiota and innate immunity in recurrent Clostridium difficile infection. J Immunol Res 2014, 462740, doi:10.1155/2014/462740 (2014).

20 Iacob, S., Iacob, D. G. & Luminos, L. M. Intestinal Microbiota as a Host Defense Mechanism to Infectious Threats. Front Microbiol 9, 3328, doi:10.3389/fmicb.2018.03328 (2018).

21 Madan, R. & Petri, W. A., Jr. Immune responses to Clostridium difficile infection. Trends Mol Med 18, 658–666, doi:10.1016/j.molmed.2012.09.005 (2012).

22 Sun, X., Savidge, T. & Feng, H. The enterotoxicity of Clostridium difficile toxins. Toxins (Basel) 2, 1848–1880, doi:10.3390/toxins2071848 (2010).

23 Kyne, L., Warny, M., Qamar, A. & Kelly, C. P. Association between antibody response to toxin A and protection against recurrent Clostridium difficile diarrhoea. Lancet 357, 189–193, doi:10.1016/S0140-6736(00)03592-3 (2001).

24 Wilcox, M. H. et al. Bezlotoxumab for Prevention of Recurrent Clostridium difficile Infection. N Engl J Med 376, 305–317, doi:10.1056/NEJMoa1602615 (2017).

25 Giannasca, P. J. et al. Serum antitoxin antibodies mediate systemic and mucosal protection from Clostridium difficile disease in hamsters. Infect Immun 67, 527–538 (1999).

26 Johnston, P. F., Gerding, D. N. & Knight, K. L. Protection from Clostridium difficile infection in CD4 T Cell- and polymeric immunoglobulin receptor-deficient mice. Infect Immun 82, 522–531, doi:10.1128/IAI.01273-13 (2014).

27 Rupnik, M., Wilcox, M. H. & Gerding, D. N. Clostridium difficile infection: new developments in epidemiology and pathogenesis. Nat Rev Microbiol 7, 526–536, doi: 10.1038/nrmicro2164 (2009).

28 Schaffler, H. & Breitruck, A. Clostridium difficile - From Colonization to Infection. Front Microbiol 9, 646, doi:10.3389/fmicb.2018.00646 (2018).

29 Blixt, T. et al. Asymptomatic Carriers Contribute to Nosocomial Clostridium difficile Infection: A Cohort Study of 4508 Patients. Gastroenterology 152, 1031–1041 e1032, doi:10.1053/j.gastro.2016.12.035 (2017).

30 Guerrero, D. M. et al. Asymptomatic carriage of toxigenic Clostridium difficile by hospitalized patients. J Hosp Infect 85, 155–158, doi:10.1016/j.jhin.2013.07.002 (2013).

31 Tenover, F. C., Baron, E. J., Peterson, L. R. & Persing, D. H. Laboratory diagnosis of Clostridium difficile infection can molecular amplification methods move us out of uncertainty? J Mol Diagn 13, 573–582, doi:10.1016/j.jmoldx.2011.06.001 (2011).

32 Burnham, C. A. & Carroll, K. C. Diagnosis of Clostridium difficile infection: an ongoing conundrum for clinicians and for clinical laboratories. Clin Microbiol Rev 26, 604–630, doi:10.1128/CMR.00016-13 (2013).

33 Musher, D. M. et al. Detection of Clostridium difficile toxin: comparison of enzyme immunoassay results with results obtained by cytotoxicity assay. J Clin Microbiol 45, 2737–2739, doi:10.1128/JCM.00686-07 (2007).

34 Kociolek, L. K. et al. Impact of a Healthcare Provider Educational Intervention on Frequency of Clostridium difficile Polymerase Chain Reaction Testing in Children: A Segmented Regression Analysis. J Pediatric Infect Dis Soc 6, 142–148, doi: 10.1093/jpids/piw027 (2017).

35 Abos, A. et al. Discriminating cognitive status in Parkinson’s disease through functional connectomics and machine learning. Sci Rep 7, 45347, doi:10.1038/srep45347 (2017).

36 Dagliati, A. et al. Machine Learning Methods to Predict Diabetes Complications. J Diabetes Sci Technol 12, 295–302, doi:10.1177/1932296817706375 (2018).

37 Mossotto, E. et al. Classification of Paediatric Inflammatory Bowel Disease using Machine Learning. Sci Rep 7, 2427, doi:10.1038/s41598-017-02606-2 (2017).

38 Kim, S. J., Cho, K. J. & Oh, S. Development of machine learning models for diagnosis of glaucoma. PLoS One 12, e0177726, doi:10.1371/journal.pone.0177726 (2017).

39 Kelly, C. P. et al. Host Immune Markers Distinguish Clostridioides difficile Infection From Asymptomatic Carriage and Non-C. difficile Diarrhea. Clin Infect Dis, doi:10.1093/cid/ciz330 (2019).

40 Mandal, S. et al. Analysis of composition of microbiomes: a novel method for studying microbial composition. Microb Ecol Health Dis 26, 27663, doi:10.3402/mehd.v26.27663 (2015).

41 Friedman, J. & Alm, E. J. Inferring correlation networks from genomic survey data. PLoS Comput Biol 8, e1002687, doi:10.1371/journal.pcbi.1002687 (2012).

42 Kuntal, B. K., Chandrakar, P., Sadhu, S. & Mande, S. S. ‘NetShift’: a methodology for understanding ‘driver microbes’ from healthy and disease microbiome datasets. ISME J 13, 442–454, doi:10.1038/s41396-018-0291-x (2019).

43 Bannister, C. A., Halcox, J. P., Currie, C. J., Preece, A. & Spasic, I. A genetic programming approach to development of clinical prediction models: A case study in symptomatic cardiovascular disease. PLoS One 13, e0202685, doi:10.1371/journal.pone.0202685 (2018).

44 Schmidt, M. & Lipson, H. Distilling free-form natural laws from experimental data. Science 324, 81–85, doi:10.1126/science.1165893 (2009).

45 Gateau, C., Couturier, J., Coia, J. & Barbut, F. How to: diagnose infection caused by Clostridium difficile. Clin Microbiol Infect 24, 463–468, doi:10.1016/j.cmi.2017.12.005 (2018).

46 Loreau, M. et al. Biodiversity and ecosystem functioning: current knowledge and future challenges. Science 294, 804–808, doi:10.1126/science.1064088 (2001).

47 Ives, A. R. & Carpenter, S. R. Stability and diversity of ecosystems. Science 317, 58–62, doi:10.1126/science.1133258 (2007).

48 Ma, Z. S., Li, L. & Gotelli, N. J. Diversity-disease relationships and shared species analyses for human microbiome-associated diseases. ISME J 13, 1911–1919, doi:10.1038/s41396-019-0395-y (2019).

49 Song, Y. et al. Microbiota dynamics in patients treated with fecal microbiota transplantation for recurrent Clostridium difficile infection. PLoS One 8, e81330, doi:10.1371/journal.pone.0081330 (2013).

50 Milani, C. et al. Gut microbiota composition and Clostridium difficile infection in hospitalized elderly individuals: a metagenomic study. Sci Rep 6, 25945, doi:10.1038/srep25945 (2016).

51 Jiang, Z. D. et al. Randomised clinical trial: faecal microbiota transplantation for recurrent Clostridum difficile infection - fresh, or frozen, or lyophilised microbiota from a small pool of healthy donors delivered by colonoscopy. Aliment Pharmacol Ther 45, 899–908, doi:10.1111/apt.13969 (2017).

52 Shankar, V. et al. Species and genus level resolution analysis of gut microbiota in Clostridium difficile patients following fecal microbiota transplantation. Microbiome 2, 13, doi:10.1186/2049-2618-2-13 (2014).

53 Zaneveld, J. R., McMinds, R. & Vega Thurber, R. Stress and stability: applying the Anna Karenina principle to animal microbiomes. Nat Microbiol 2, 17121, doi:10.1038/nmicrobiol.2017.121 (2017).

54 Giongo, A. et al. Toward defining the autoimmune microbiome for type 1 diabetes. J 5, 82–91, doi:10.1038/ismej.2010.92 (2011).

55 Caussy, C. et al. A gut microbiome signature for cirrhosis due to nonalcoholic fatty liver disease. Nat Commun 10, 1406, doi:10.1038/s41467-019-09455-9 (2019).

56 Rowan, F. et al. Desulfovibrio bacterial species are increased in ulcerative colitis. Dis Colon Rectum 53, 1530–1536, doi:10.1007/DCR.0b013e3181f1e620 (2010).

57 Kolling, G. L. et al. Lactic acid production by Streptococcus thermophilus alters Clostridium difficile infection and in vitro Toxin A production. Gut Microbes 3, 523–529, doi:10.4161/gmic.21757 (2012).

58 van de Beek, D. et al. Clinical features and prognostic factors in adults with bacterial meningitis. N Engl J Med 351, 1849–1859, doi:10.1056/NEJMoa040845 (2004).

59 Arnold, R. S. et al. Emergence of Klebsiella pneumoniae carbapenemase-producing bacteria. South Med J 104, 40–45, doi:10.1097/SMJ.0b013e3181fd7d5a (2011).

60 Navon-Venezia, S., Kondratyeva, K. & Carattoli, A. Klebsiella pneumoniae: a major worldwide source and shuttle for antibiotic resistance. FEMS Microbiol Rev 41, 252–275, doi:10.1093/femsre/fux013 (2017).

61 Cohen, S. H. et al. Clinical practice guidelines for Clostridium difficile infection in adults: 2010 update by the society for healthcare epidemiology of America (SHEA) and the infectious diseases society of America (IDSA). Infect Control Hosp Epidemiol 31, 431–455, doi:10.1086/651706 (2010).

62 Pollock, N. R. et al. Comparison of Clostridioides difficile Stool Toxin Concentrations in Adults With Symptomatic Infection and Asymptomatic Carriage Using an Ultrasensitive Quantitative Immunoassay. Clin Infect Dis 68, 78–86, doi:10.1093/cid/ciy415 (2019).

63 Crobach, M. J. T. et al. Understanding Clostridium difficile Colonization. Clin Microbiol Rev 31, doi:10.1128/CMR.00021-17 (2018).

64 Antharam, V. C. et al. An Integrated Metabolomic and Microbiome Analysis Identified Specific Gut Microbiota Associated with Fecal Cholesterol and Coprostanol in Clostridium difficile Infection. PLoS One 11, e0148824, doi:10.1371/journal.pone.0148824 (2016).

65 Khanna, S. et al. Gut microbiome predictors of treatment response and recurrence in primary Clostridium difficile infection. Aliment Pharmacol Ther 44, 715–727, doi:10.1111/apt.13750 (2016).

66 Han, S. H., Yi, J., Kim, J. H., Lee, S. & Moon, H. W. Composition of gut microbiota in patients with toxigenic Clostridioides (Clostridium) difficile: Comparison between subgroups according to clinical criteria and toxin gene load. PLoS One 14, e0212626, doi: 10.1371/journal.pone.0212626 (2019).

67 Daquigan, N., Seekatz, A. M., Greathouse, K. L., Young, V. B. & White, J. R. High resolution profiling of the gut microbiome reveals the extent of Clostridium difficile burden. NPJ Biofilms Microbiomes 3, 35, doi:10.1038/s41522-017-0043-0 (2017).

68 Hudson, L. E., Anderson, S. E., Corbett, A. H. & Lamb, T. J. Gleaning Insights from Fecal Microbiota Transplantation and Probiotic Studies for the Rational Design of Combination Microbial Therapies. Clin Microbiol Rev 30, 191–231, doi:10.1128/CMR.00049-16 (2017).

69 Fujitani, S., George, W. L., Morgan, M. A., Nichols, S. & Murthy, A. R. Implications for vancomycin-resistant Enterococcus colonization associated with Clostridium difficile infections. Am J Infect Control 39, 188–193, doi:10.1016/j.ajic.2010.10.024 (2011).

70 Antharam, V. C. et al. Intestinal dysbiosis and depletion of butyrogenic bacteria in Clostridium difficile infection and nosocomial diarrhea. J Clin Microbiol 51, 2884–2892, doi:10.1128/JCM.00845-13 (2013).

71 Sokol, H. et al. Specificities of the intestinal microbiota in patients with inflammatory bowel disease and Clostridium difficile infection. Gut Microbes 9, 55–60, doi:10.1080/19490976.2017.1361092 (2018).

72 Mezzatesta, M. L., Gona, F. & Stefani, S. Enterobacter cloacae complex: clinical impact and emerging antibiotic resistance. Future Microbiol 7, 887–902, doi:10.2217/fmb.12.61 (2012).

73 Umana, A. et al. Utilizing Whole Fusobacterium Genomes To Identify, Correct, and Characterize Potential Virulence Protein Families. J Bacteriol 201, doi:10.1128/JB.00273-19 (2019).

74 Deng, H. Interpreting tree ensembles with in Trees. International Journal of Data Science and Analytics 7, 277–287, doi:10.1007/s41060-018-0144-8 (2019).

75 Dreiseitl, S. & Ohno-Machado, L. Logistic regression and artificial neural network classification models: a methodology review. J Biomed Inform 35, 352–359, doi:10.1016/s1532-0464(03)00034-0 (2002).

76 Tollenaar, N. & van der Heijden, P. G. M. Optimizing predictive performance of criminal recidivism models using registration data with binary and survival outcomes. PLoS One 14, e0213245, doi:10.1371/journal.pone.0213245 (2019).

77 Norton, E. C. & Dowd, B. E. Log Odds and the Interpretation of Logit Models. Health Serv Res 53, 859–878, doi:10.1111/1475-6773.12712 (2018).

78 Liu, K. H. & Xu, C. G. A genetic programming-based approach to the classification of multiclass microarray datasets. Bioinformatics 25, 331–337, doi: 10.1093/bioinformatics/btn644 (2009).

79 Fadrosh, D. W. et al. An improved dual-indexing approach for multiplexed 16S rRNA gene sequencing on the Illumina MiSeq platform. Microbiome 2, 6, doi:10.1186/2049-2618-2-6 (2014).

80 Biesheuvel, C. J., Siccama, I., Grobbee, D. E. & Moons, K. G. Genetic programming outperformed multivariable logistic regression in diagnosing pulmonary embolism. J Clin Epidemiol 57, 551–560, doi:10.1016/j.jclinepi.2003.10.011 (2004).

81 Staats, K., Pantridge, E., Cavaglia, M., Milovanov, I. & Aniyan, A. TensorFlow Enabled Genetic Programming. *arXiv e-prints*, arXiv:1708.03157 (2017).

